# Examining disparities relating to service reach and patient engagement with COVID-19 remote home monitoring services in England: a mixed methods rapid evaluation

**DOI:** 10.1101/2022.02.21.22270793

**Authors:** Nadia E Crellin, Lauren Herlitz, Manbinder S Sidhu, Jo Ellins, Theo Georghiou, Ian Litchfield, Efthalia Massou, Pei Li Ng, Chris Sherlaw-Johnson, Sonila M Tomini, Cecilia Vindrola-Padros, Holly Walton, Naomi J Fulop

## Abstract

**Background:** The adoption of remote methods of care has been accelerated by the COVID-19 pandemic, but concerns exist relating to the potential impact on health disparities. This evaluation explores the implementation of COVID-19 remote home monitoring services across England, focussing on patients’ experiences and engagement with the service.

**Methods:** The study was a rapid, multi-site, mixed methods evaluation. Data were collected between January and June 2021. We conducted qualitative interviews with staff service leads, and patients and carers receiving the service. We conducted quantitative surveys with staff delivering the service, and patients and carers receiving the service across 28 sites in England, UK. Qualitative data were analysed using thematic analysis and quantitative data were analysed using univariate and multivariate methods.

**Findings:** Many sites designed their service to be inclusive to the needs of their local population. Strategies included widening eligibility criteria, prioritising vulnerable groups, and creating referral pathways. Many sites also adapted their services according to patient needs, including providing information in different languages or more accessible formats, offering translation services, offering non-digital options, or providing face-to-face assessments. Despite these adaptions, disparities were reported across patient groups (e.g. age, health status, ethnicity, level of education) in their experience of and engagement with the service.

**Interpretation:** Services must determine how best to design and implement remote monitoring services to be of value to all populations. National guidance should play a role in supporting services to best serve the needs of their populations, and patients and staff must play an active role in service design.

**Funding:** This is independent research funded by the National Institute for Health Research, Health Services & Delivery Research programme (RSET Project no. 16/138/17; BRACE Project no. 16/138/31) and NHSEI. NJF is an NIHR Senior Investigator. The views expressed in this publication are those of the authors and not necessarily those of the National Institute for Health Research or the Department of Health and Social Care.

**Research in context:** *Evidence before this study:* Evidence shows COVID-19 has a disproportionate impact on certain population groups, such as ethnic minority groups, older adults and those with comorbidities. The rapid adoption and spread of remote home monitoring services in England must be accompanied by evaluations at a local level to monitor the impact on health disparities in local populations.

*Added value of this study:* This rapid mixed methods evaluation of COVID-19 home monitoring services adopted across 28 sites in England aimed to increase understanding of how services have been designed and delivered to address local population needs to increase accessibility to the service and facilitate engagement with the service. We add to the literature by identifying a range of local service adaptations which aim to increase reach and facilitate patient engagement, and consider their potential impact on health disparities. We found strategies included prioritising vulnerable groups, creating referral pathways, offering translation services, offering non-digital options, or providing face-to-face assessments. Despite efforts to adapt services to meet local needs, disparities across patient groups in their experience of, and engagement with, the service (related to age, health status, ethnicity, and level of education) were reported.

*Implications of the available evidence:* At both a national and local level, and particularly given the increasing use of remote home monitoring schemes, lessening health disparities must be a primary focus in the design and delivery of remote monitoring models for COVID-19 and other conditions. Future research should focus on how best to design and evaluate remote monitoring services, for a range of conditions, especially for patients residing in areas where significant health disparities persist, as well as addressing the effectiveness of any strategies on specific population groups.

## Introduction

Since the start of the pandemic, remote home monitoring models for COVID-19 patients have been implemented in several countries^1^. These services look to ensure patients are escalated earlier to avoid invasive ventilation, intensive care admission, to reduce unnecessary attendance at emergency departments and transmission of the virus.

Remote home monitoring services have previously been implemented in the UK for chronic conditions^2,3^, and during the first wave of the pandemic these services were set-up and piloted for COVID-19 patients in England^4^. These services were subsequently rolled out nationally, known as ‘COVID Oximetry at home (CO@h)’ or ‘Virtual Wards’ for COVID-19 patients who had been discharged early from hospital. Patients using these services were provided with a pulse oximeter and asked to measure oxygen saturation levels, and take other measurements to enable the remote assessment of their condition. Many sites adopted technology-enabled methods, for example, using digital applications, web links, or automated texts/calls, alongside analogue methods (i.e. telephone calls) to submit readings to the service^4^.

The COVID-19 pandemic has shone a spotlight on existing health disparities in the UK. The pandemic has disproportionately affected the most marginalised communities; particularly people living in more socio-economically deprived areas, those from minority ethnic communities, older adults, people with a learning disability and those residing in care homes^5,6^. The pandemic has also led to changes to the way that health services are delivered at an unprecedented pace^7,8^, with a rapid adoption of digital technology and an acceleration of the move towards remote monitoring models of care. Little is known whether and how this shift might affect existing health disparities^9–11^. Inequalities can be introduced at all stages of the planning and delivery of health interventions, including: access to the intervention, diagnostic accuracy, patient uptake and adherence^12^. Addressing inequalities in health care access, quality, use and outcomes is high on the health policy agenda of many countries, and a key focus of the NHS long term plan^13^.

The benefits brought about by remote methods of delivering health care are not experienced by all. In the case of tech-enabled remote methods, digital exclusion may negatively impact access to or quality of care^14,15^. For example, older adults are disproportionately affected by COVID-19, yet are more likely to lack digitals skills and infrastructure^16^. Other factors beyond technology may exacerbate disparities in access or engagement with remote monitoring; some patients may need additional support to enable them to engage appropriately with remote health services due to lower health literacy or understanding how health systems work^17^.

Studies of the effectiveness of strategies to reduce health disparities for remote monitoring services are scarce. Two studies on patients’ access and engagement with telehealth proposed: improving digital infrastructure; collecting and monitoring data on technology access and literacy; and co-developing platforms^18,19^. In wider literature, strategies for reducing disparities in access and use of health services includes: collecting and analysing data on relevant populations; redistributing/targeting resources in the population; collaborating with communities where health disparities are known to exist; strategies to reach high-risk populations (e.g. pop-up services in community spaces); using culturally and linguistically appropriate materials; and training health professionals to share best practice^20–23^. The effectiveness of strategies, however, is largely unknown due to differences in reporting on health inequalities and a lack of high quality studies^24^.

Given the recent rapid shift towards remote models of care, it is therefore, important to explore whether disparities exist in how patients access, engage with and experience services, and to understand whether and how remote services can be designed and delivered to promote inclusion for all populations. This evaluation of remote home monitoring models in England addressed the following questions:

1. Were COVID-19 remote home monitoring services adapted at a local level to increase service reach and patient engagement?
2. Were there disparities in patients’ reports of their ability to engage with the service, and were these moderated by the modality of the service? What were the potential impacts of service adaptations on health disparities according to patients?
3. Were there disparities in patients’ reports of their experience of the service?

## Methods

### Study design

This multi-site, mixed methods study was part of a larger rapid evaluation of remote COVID-19 home monitoring services^25^. The evaluation was carried out in England, UK between January and June 2021. The methods for this study are reported in detail in Appendix 1.

### Conceptual framework – National Institute on Aging (NIA) Health Disparities Research Framework

We drew upon the National Institute on Aging (NIA) health disparities research framework to develop our research questions and inform our analysis^26^. This framework was selected as it highlights a comprehensive range of factors (i.e. environmental, sociocultural, behavioural and biological) that determine priority populations for health disparities research related to aging. This was considered relevant to this programme as older adults were the primary target population of COVID-19 remote monitoring services. The framework identifies fundamental factors as important to all levels of analysis. We focused our analysis on most of these factors, including gender, age, ethnicity, socioeconomic status, disability status, and two key environmental factor sub-domains– socioeconomic and geographic factors – to examine disparities in relation to access, experience and engagement. We considered two additional factors: living situation (socio-economic factor) service and deprivation score (i.e. deciles of the Index of Multiple Deprivation) as a geographic factor.

### Sampling and data collection

Twenty-eight sites were selected using a range of criteria (i.e. setting, type of model, mode of monitoring). Seventeen of the 28 sites were selected as case study sites for more in-depth qualitative analysis.

#### Quantitative surveys

We asked service leads to complete an online survey, and to distribute the survey link to staff involved in service delivery. In the survey we asked staff to identify any patient groups facing barriers to engagement and whether the service had been adapted locally to address any specific patient needs. Service leads coordinated the distribution of the patient survey (25/28 sites). Patients or carers received an electronic or paper version. The survey focused on patients’ experiences of and engagement with the service including understanding the information provided, completion of tasks, whether they encountered problems, and whether they would make any changes to the service (see Appendix 2 for staff and patient survey questions).

#### Qualitative interviews

We carried out semi-structured interviews with 23 service delivery leads from 16 of the 17 case study sites. Interviews were carried out over the phone or via MS Teams. Interviews focused on the design and implementation of services, and staff experiences of implementation, including barriers and enablers.

Service leads from the 17 case study sites were asked to identify four to six patients to invite to participate in the study. Patient interviews focused on: how patients were referred to the service, how they felt about recording and monitoring their symptoms, how they communicated with the service and their experience of escalating care (see Appendix 3 for topic guides).

An informed consent process using participant information sheets and written consent was used for both staff and patient interviews to ensure informed and voluntary participation.

### Data analysis

#### Quantitative surveys

We analysed survey data using SPSS statistical software (version 25). Descriptive statistics were calculated to explore staff perceptions of patient groups facing barriers and the number of services that made adaptations according to patient needs. Open text responses relating to local service adaptations and patient groups facing barriers were analysed, triangulated with service lead interview data, and coded into themes (see Table 3 in results).

For patient survey data, we used descriptive statistics. Non-parametric univariate analyses were conducted to explore patient engagement with the service across fundamental factors; age, gender, ethnicity, and health status, socio-economic factors; education, employment status, English as first language, and living situation, and the geographic factor; deprivation score. We used Mann-Whitney U tests and Kruskal-Wallis H tests. Due to the large number of statistical hypothesis tests conducted, and hence the possibility of false positive results at the traditional level of statistical significance of p<0.05, it was determined that a level of p<0.01 should be used. Similar non-parametric univariate analyses were used to explore patient experiences across the same fundamental, socio-economic and geographic factors.

We conducted logistic regression modelling to examine whether modality of the service, age, education, health status and ethnicity were associated with likelihood of patients reporting a problem with the service. Patient open text survey responses providing feedback about the service were triangulated with patient interview data and coded into themes related to service design and engagement barriers.

#### Service lead and patient interviews

To understand whether and how local services were adapted to address health disparities, a thematic analysis was carried out on service lead interview data^27,28^. Coding was conducted using NVivo 12 software and organised into themes related to service adaptations aimed at increasing reach or engagement (see Table 3 in results).

To examine the potential impacts of adaptations from patients’ perspectives, rapid assessment procedure (RAP) sheets were analysed and data was deductively coded using the framework outlined by the themes from the service lead interviews^29^.

## Results

Twenty-eight sites across England were included in the evaluation (see Table 1 for site characteristics).

**Table 1.** Summary of site characteristics

|  | Site characteristic/domain | Number of sites<br>(n=28) | Number of case study<br>sites (n=17) |
| --- | --- | --- | --- |
| Type of model | CO@H<sup>a</sup> | 13 | 9 |
|  | Virtual ward<sup>b</sup> | 4 | 1 |
|  | Integrated CO@H and virtual ward | 11 | 7 |
| Sector leading the<br>service | Primary care/community care | 16 | 11 |
|  | Secondary care | 5 | 3 |
|  | Both | 5 | 3 |
|  | Not specified | 2 | 0 |
| Mode of patient<br>monitoring | Analogue only<sup>c</sup> | 7 | 3 |
|  | Tech-enabled and analogue<sup>d</sup> | 21 | 14 |
| Geographic location | South West England | 7 | 3 |
|  | South East England | 5 | 4 |
|  | East of England | 1 | 1 |
|  | Greater London | 5 | 3 |
|  | East Midlands | 2 | 1 |
|  | North East England | 2 | 1 |
|  | North West England | 5 | 4 |
|  | Yorkshire and Humber | 1 | 0 |
| Month service started | Before November 2020 | 11 | 7 |
|  | In November 2020 | 13 | 9 |
|  | After November 2020 | 4 | 1 |

<sup>a</sup>Pre-hospital model; <sup>b</sup>Early discharge from hospital model <sup>c</sup>Paper and telephone; <sup>d</sup>A mixture of phone call and tech-enabled methods (i.e. app, web link)

### Participant characteristics

We interviewed 23 service delivery leads and 62 patients and carers (see Appendix 5 for interviewee demographics).

We received 292 staff surveys across 28 sites (70 service managers/leads and 222 delivery staff; 39% response rate). See Appendix 6 for staff survey respondent characteristics. We received 1069 surveys (18% response rate) from patients and carers across 25 sites. Of the total respondents, 97.6% (n=936/1069) were patients, 4.5% (n=48/1069) were carers and for 8% (n=85/1069) of respondents it was not possible to determine if a patient or carer completed the survey. Table 2 presents patient survey respondent characteristics.

**Table 2.** Demographic characteristics for patient and carer survey respondents

| Demographic characteristic |  | Patient<br>N (%) | Carer<br>N (%) |
| --- | --- | --- | --- |
| Gender<br>(patient N=920; carer N=45) | Female | 531 (58) | 27 (60) |
|  | Male | 385 (42) | 18 (40) |
|  | Other/prefer not to say | 4 (0.4) | 0 |
| Age<br>(patient N=923; carer N=46) | Under 50 years | 195 (21.1) | 13 (28.3) |
|  | 50-64 years | 428 (46.4) | 24 (52.2) |
|  | 65-79 years | 256 (27.8) | 4 (8.7) |
|  | >=80 years | 43 (4.7) | 5 (10.9) |
|  | Prefer not to say | 1 (0.1) | 0 |
| Living situation (patient N=863) | Living alone | 132 (15.3) |  |
|  | Household of 2 | 339 (39.3) |  |
|  | Household of 3 | 152 (17.6) |  |
|  | Household of 4/5/ | 201 (23.3) |  |
|  | Household of 6+ | 36 (4.2) |  |
|  | Prefer not to say | 3 (0.3) |  |
| Ethnicity<br>(patient N=918; carer N=47) | White British or any other White background | 836 (91.1) | 38 (80.9) |
|  | Black/African/Caribbean/Black British or any other Black background | 16 (1.7) | 0 |
|  | Asian/Asian British or any other Asian background | 48 (5.2) | 9 (19.1) |
|  | Mixed or multiple ethnic background | 12 (1.3) | 0 |
|  | Any other ethnic group | 2 (0.2) | 0 |
|  | Prefer not to say | 4 (0.4) | 0 |
| Highest educational qualification<br>(patient N=914; carer N=46) | No formal qualification | 146 (16) | 10 (21.7) |
|  | GCSE/CSE/O level or equivalent | 273 (29.9) | 16 (34.8) |
|  | A level/AS level or equivalent | 106 (11.6) | 8 (17.4) |
|  | Degree level or higher | 212 (23.2) | 7 (15.2) |
|  | Other | 80 (8.8) | 1 (2.2) |
|  | Prefer not to say/not sure | 97 (10.6) | 4 (8.7) |
| Work situation*<br>(patient N=969; carer N=48) | Working full time | 355 (37.6) | 16 (33.3) |
|  | Working part time | 128 (13.5) | 6 (12.5) |
|  | Self-employed | 41 (4.3) | 1 (2.1) |
|  | Student in higher education | 2 (0.2) | 0 |
|  | Unemployed | 18 (1.9) | 3 (6.3) |
|  | Homemaker | 21 (2.2) | 3 (6.3) |
|  | Retired | 274 (29) | 9 (18.8) |
|  | Furloughed | 15 (1.6) | 0 |
|  | Full time carer | 19 (2) | 1 (2.1) |
|  | Not in work due to poor health or disability | 65 (6.9) | 5 (10.4) |
|  | Other/prefer not to say | 31 (3.3) | 1 (2.1) |
| English as first language (patient N=925; carer N=43) | Yes | 852 (92.1) | 35 (81.4) |
|  | No | 66 (7.1) | 8 (18.6) |
|  | Prefer not to say | 7 (0.8) | 0 |
| Day to day activities limited by a health problem or disability<br>(patient N=920; carer N=46) | Limited a lot or a little | 351 (38.1) | 20 (43.4) |
|  | Not limited at all | 482 (52.4) | 17 (37) |
|  | Prefer not to say | 4 (0.4) | 1 (2.2) |
|  | Not sure/not applicable | 83 (9) | 8 (17.4) |
| Deprivation score**<br>(patient N=767; carer N=37) | D1 or 2 (Most deprived) | 182 (23.7) | 13 (35.1) |
|  | D3 or 4 | 137 (17.9) | 5 (13.5) |
|  | D5 or 6 | 149 (19.4) | 7 (18.9) |
|  | D7 or 8 | 161 (21) | 9 (24.3) |
|  | D9 or 10 (Least deprived) | 18 (18) | 3 (8.1) |

\*Respondents able to select more than one option
\*\* Deprivation by LSOA (Index of Multiple Deprivation decile)

**Table 3.**
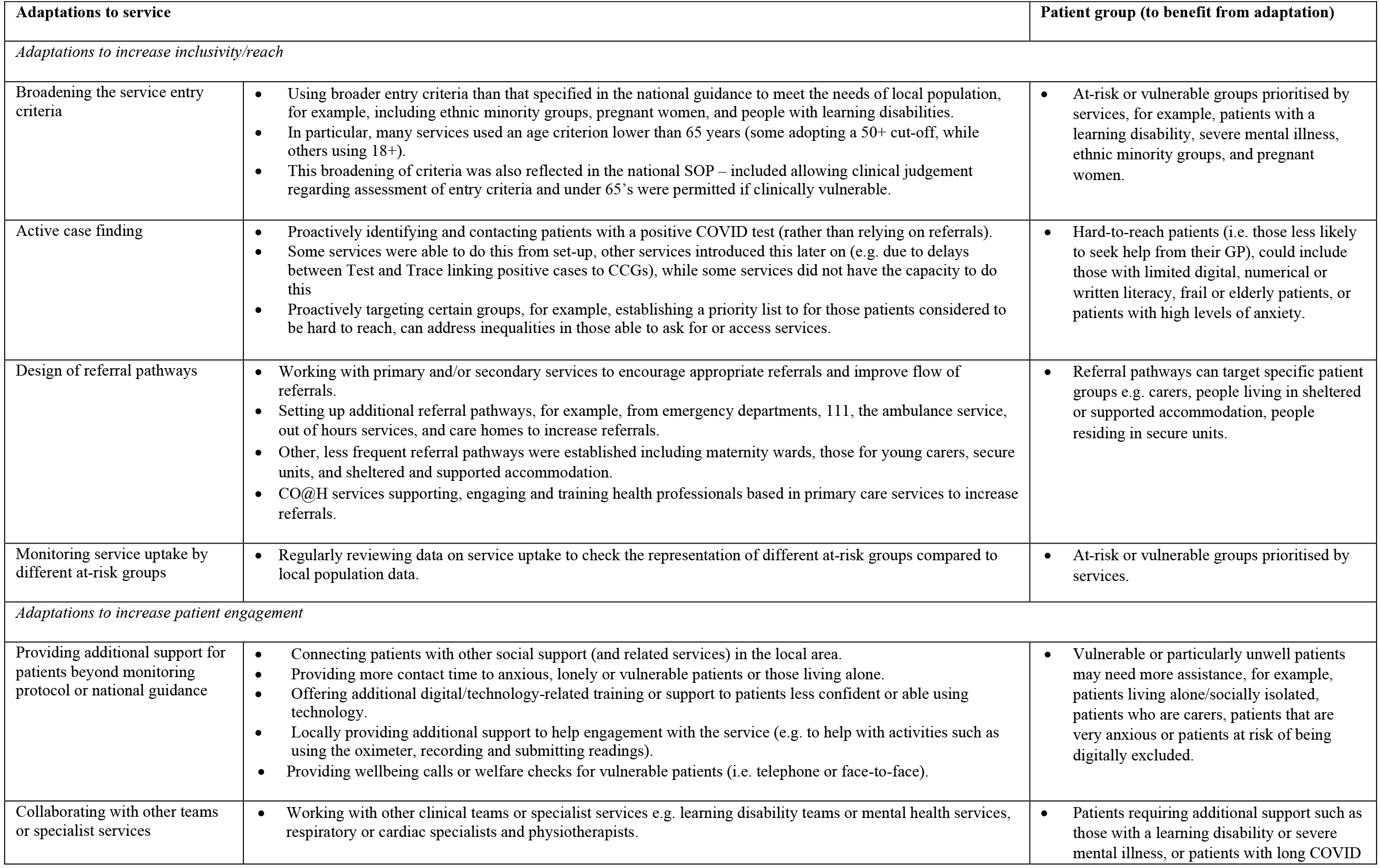
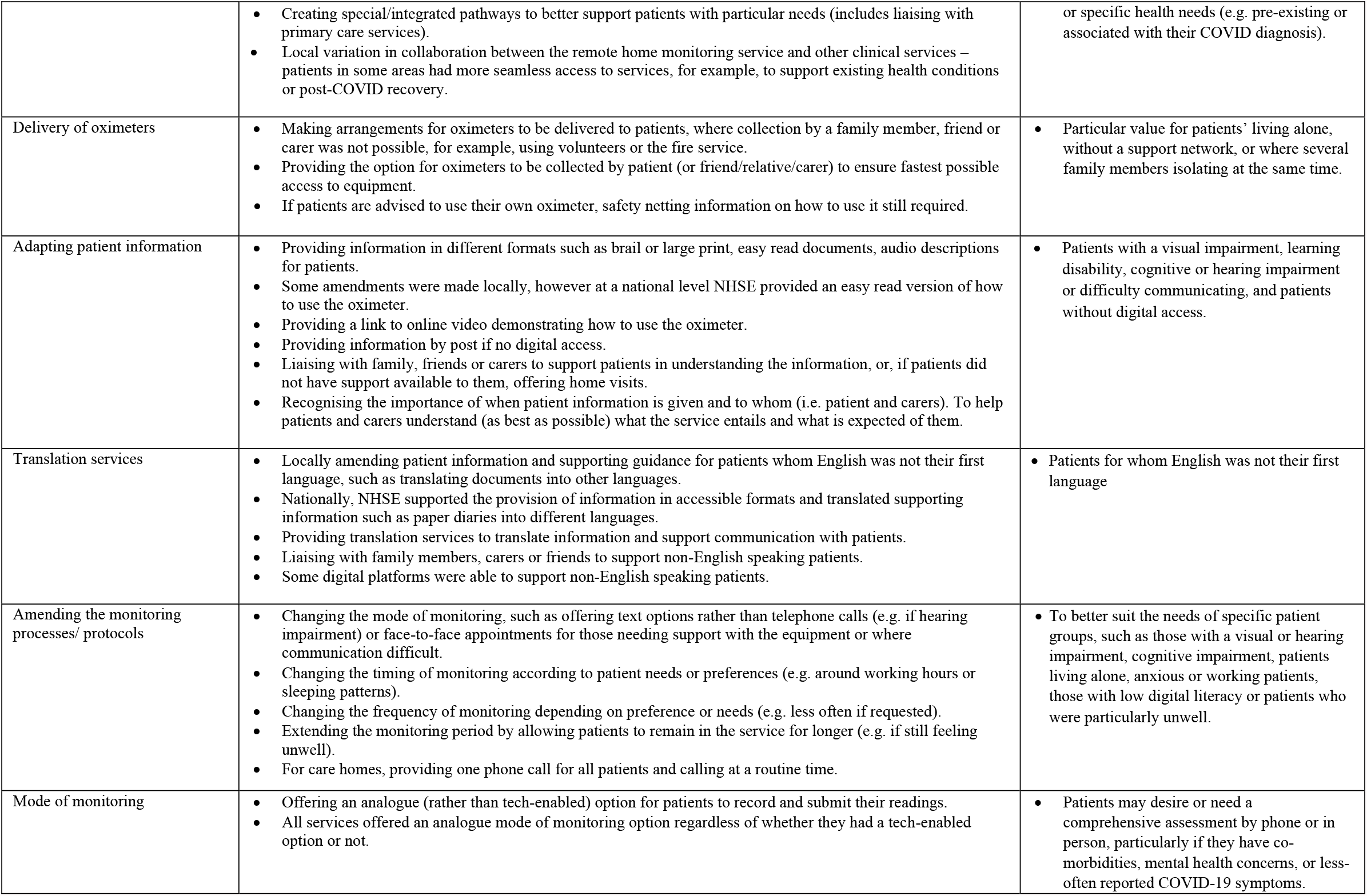
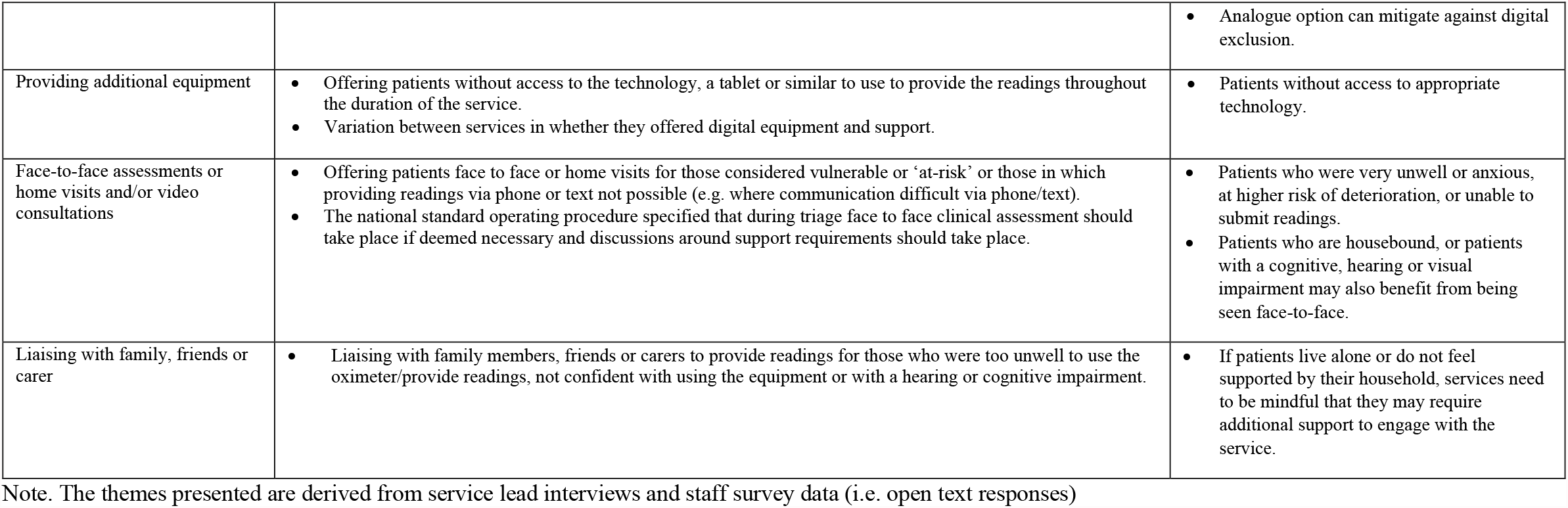
Local service adaptations to increase reach and facilitate patient engagement

| Adaptations to service |  | Patient group (to benefit from adaptation) |
| --- | --- | --- |
| *Adaptations to increase inclusivity/reach* |  |  |
| Broadening the service entry criteria | Using broader entry criteria than that specified in the national guidance to meet the needs of local population, for example, including ethnic minority groups, pregnant women, and people with learning disabilities.<br>In particular, many services used an age criterion lower than 65 years (some adopting a 50+ cut-off, while others using 18+).<br>This broadening of criteria was also reflected in the national SOP – included allowing clinical judgement regarding assessment of entry criteria and under 65's were permitted if clinically vulnerable. | At-risk or vulnerable groups prioritised by services, for example, patients with a learning disability, severe mental illness, ethnic minority groups, and pregnant women. |
| Active case finding | Proactively identifying and contacting patients with a positive COVID test (rather than relying on referrals).<br>Some services were able to do this from set-up, other services introduced this later on (e.g. due to delays between Test and Trace linking positive cases to CCGs), while some services did not have the capacity to do this<br>Proactively targeting certain groups, for example, establishing a priority list to for those patients considered to be hard to reach, can address inequalities in those able to ask for or access services. | Hard-to-reach patients (i.e. those less likely to seek help from their GP), could include those with limited digital, numerical or written literacy, frail or elderly patients, or patients with high levels of anxiety. |
| Design of referral pathways | Working with primary and/or secondary services to encourage appropriate referrals and improve flow of referrals.<br>Setting up additional referral pathways, for example, from emergency departments, 111, the ambulance service, out of hours services, and care homes to increase referrals.<br>Other, less frequent referral pathways were established including maternity wards, those for young carers, secure units, and sheltered and supported accommodation.<br>CO@H services supporting, engaging and training health professionals based in primary care services to increase referrals. | Referral pathways can target specific patient groups e.g. carers, people living in sheltered or supported accommodation, people residing in secure units. |
| Monitoring service uptake by different at-risk groups | Regularly reviewing data on service uptake to check the representation of different at-risk groups compared to local population data. | At-risk or vulnerable groups prioritised by services. |
| *Adaptations to increase patient engagement* |  |  |
| Providing additional support for patients beyond monitoring protocol or national guidance | Connecting patients with other social support (and related services) in the local area.<br>Providing more contact time to anxious, lonely or vulnerable patients or those living alone.<br>Offering additional digital/technology-related training or support to patients less confident or able using technology.<br>Locally providing additional support to help engagement with the service (e.g. to help with activities such as using the oximeter, recording and submitting readings).<br>Providing wellbeing calls or welfare checks for vulnerable patients (i.e. telephone or face-to-face). | Vulnerable or particularly unwell patients may need more assistance, for example, patients living alone/socially isolated, patients who are carers, patients that are very anxious or patients at risk of being digitally excluded. |
| Collaborating with other teams or specialist services | Working with other clinical teams or specialist services e.g. learning disability teams or mental health services, respiratory or cardiac specialists and physiotherapists. | Patients requiring additional support such as those with a learning disability or severe mental illness, or patients with long COVID |
|  | • Creating special/integrated pathways to better support patients with particular needs (includes liaising with primary care services).<br>• Local variation in collaboration between the remote home monitoring service and other clinical services – patients in some areas had more seamless access to services, for example, to support existing health conditions or post-COVID recovery. | or specific health needs (e.g. pre-existing or associated with their COVID diagnosis). |
| Delivery of oximeters | • Making arrangements for oximeters to be delivered to patients, where collection by a family member, friend or carer was not possible, for example, using volunteers or the fire service.<br>• Providing the option for oximeters to be collected by patient (or friend/relative/carers) to ensure fastest possible access to equipment.<br>• If patients are advised to use their own oximeter, safety netting information on how to use it still required. | • Particular value for patients' living alone, without a support network, or where several family members isolating at the same time. |
| Adapting patient information | • Providing information in different formats such as brail or large print, easy read documents, audio descriptions for patients.<br>• Some amendments were made locally, however at a national level NHSE provided an easy read version of how to use the oximeter.<br>• Providing a link to online video demonstrating how to use the oximeter.<br>• Providing information by post if no digital access.<br>• Liaising with family, friends or carers to support patients in understanding the information, or, if patients did not have support available to them, offering home visits.<br>• Recognising the importance of when patient information is given and to whom (i.e. patient and carers). To help patients and carers understand (as best as possible) what the service entails and what is expected of them. | • Patients with a visual impairment, learning disability, cognitive or hearing impairment or difficulty communicating, and patients without digital access. |
| Translation services | • Locally amending patient information and supporting guidance for patients whom English was not their first language, such as translating documents into other languages.<br>• Nationally, NHSE supported the provision of information in accessible formats and translated supporting information such as paper diaries into different languages.<br>• Providing translation services to translate information and support communication with patients.<br>• Liaising with family members, carers or friends to support non-English speaking patients.<br>• Some digital platforms were able to support non-English speaking patients. | • Patients for whom English was not their first language |
| Amending the monitoring processes/ protocols | • Changing the mode of monitoring, such as offering text options rather than telephone calls (e.g. if hearing impairment) or face-to-face appointments for those needing support with the equipment or where communication difficult.<br>• Changing the timing of monitoring according to patient needs or preferences (e.g. around working hours or sleeping patterns).<br>• Changing the frequency of monitoring depending on preference or needs (e.g. less often if requested).<br>• Extending the monitoring period by allowing patients to remain in the service for longer (e.g. if still feeling unwell).<br>• For care homes, providing one phone call for all patients and calling at a routine time. | • To better suit the needs of specific patient groups, such as those with a visual or hearing impairment, cognitive impairment, patients living alone, anxious or working patients, those with low digital literacy or patients who were particularly unwell. |
| Mode of monitoring | • Offering an analogue (rather than tech-enabled) option for patients to record and submit their readings.<br>• All services offered an analogue mode of monitoring option regardless of whether they had a tech-enabled option or not. | • Patients may desire or need a comprehensive assessment by phone or in person, particularly if they have co-morbidities, mental health concerns, or less-often reported COVID-19 symptoms. |
|  |  | Analogue option can mitigate against digital exclusion. |
| Providing additional equipment | Offering patients without access to the technology, a tablet or similar to use to provide the readings throughout the duration of the service.<br>Variation between services in whether they offered digital equipment and support. | Patients without access to appropriate technology. |
| Face-to-face assessments or home visits and/or video consultations | Offering patients face to face or home visits for those considered vulnerable or 'at-risk' or those in which providing readings via phone or text not possible (e.g. where communication difficult via phone/text).<br>The national standard operating procedure specified that during triage face to face clinical assessment should take place if deemed necessary and discussions around support requirements should take place. | Patients who were very unwell or anxious, at higher risk of deterioration, or unable to submit readings.<br>Patients who are housebound, or patients with a cognitive, hearing or visual impairment may also benefit from being seen face-to-face. |
| Liaising with family, friends or carer | Liaising with family members, friends or carers to provide readings for those who were too unwell to use the oximeter/provide readings, not confident with using the equipment or with a hearing or cognitive impairment. | If patients live alone or do not feel supported by their household, services need to be mindful that they may require additional support to engage with the service. |

Note. The themes presented are derived from service lead interviews and staff survey data (i.e. open text responses)

Comparison of patient survey respondent characteristics with those of more than 26,000 patients onboarded to CO@h services between October 2020 and April 2021^30^ indicate the survey sample to be relatively representative of patients engaging with the service. However, several differences should be noted. The survey sample is under-representative of patients over 80 years (5% vs 10%) and under 50 years of age (21% vs 33%), and over-representative of patients aged 50-80 years (74% vs 57%). The survey sample comprises a higher proportion of patients from white ethnic groups (91%) compared to those onboarded to the service (76%), and a lower proportion of patients from minority ethnic groups. Finally, our sample is over-representative of patients in the least deprived deciles of the Index of Multiple Deprivation (18% vs 13%) and under-representative of patients in the most deprived deciles (24% vs 28%). It should be noted the survey sample includes patients onboarded to either CO@h services or virtual wards.

### Service adaptations to increase service reach and patient engagement

Staff reported their views of patient groups facing barriers to engaging with the service. Thirty-nine per cent (n=113/292) reported patients for whom English was not their first language, 33% (n=97/292) patients with a visual or hearing impairment, 31% (n=91/292) cognitive impairment, 22% (n=63/292) patients with a learning disability, 21% (n=61/292) those who are digitally excluded, 13% (n=37/292) older adults, 7% (n=20/292) ethnic minority groups, and 5% (n=16/292) unpaid carers. Other patient groups reported by staff in surveys or interviews as facing barriers included those living alone, patients unable to collect the oximeter, people with mental health problems, pregnant patients, patients whose GP had not engaged with the service, those with difficulty communicating via phone, and patients without a fixed abode.

Two-thirds (66%) of service leads for 16 of the 24 sites who provided a response reported that their service had been adapted at a local level to accommodate specific service user needs. Examples included providing information in different languages, offering translation services, offering non-digital monitoring options, face-to-face assessments, and flexibility of monitoring. Service lead interviews and survey data indicated there was considerable local variation in the number of strategies adopted by sites, for example, several services made considerable efforts to standardise the coverage of the service, invest resources into setting up pathways targeting hard-to-reach, vulnerable or at-risk groups, and to monitor uptake. Local service adaptations relating to inclusion and engagement reported by staff are set out in Table 3.

### Disparities in patients’ reports of their ability to engage with the service

In this section, we consider patient reported engagement with the service in terms of the accessibility of information, the achievability of tasks, and problems reported (drawn from patient survey data, see appendix 7 for further details). We also consider patient feedback from interviews and open-text survey responses relating to patient views and experiences of the impact of service adaptations aimed at addressing inequalities. Details relating to patient experience and engagement, more broadly, are reported elsewhere^31^.

#### Accessing the service

Patients reported being referred to the service by many different organisations and pathways (qualitative data). However, sites were not always consistent in the admission criteria applied to access the service; several patients reported inconsistencies across local areas in who was referred to the service. There was sometimes a lack of clarity in how patients were referred to the service; several patients from sites that used active case finding reported that they did not know how they were referred to the service. Collaboration between services was perceived to have facilitated more rapid access to care; several patients highlighted that they were supported quickly because of close liaison between the service and their GP practice, hospital or ambulance services. Once enrolled into the service, there were challenges relating to obtaining the appropriate equipment as part of the service; some patients reported it was difficult to collect the oximeter when they were unwell and relied on family members or friends to collect the oximeter, which might be a particular concern for patients without a support network. Many patients purchased their own oximeter or were given them by a friend or family member.

#### Accessibility of the information about the service

Qualitative interviews and open text survey responses indicated variation across and between sites in the information available to patients and how it was provided. The survey analysis found evidence of a difference in understanding information provided by the service across patient age categories (p=0.005) and ethnic groups (p=0.001). There was also some indication of differences with health status (p=0.014).

Patients aged 80 years and over reported less ease in understanding information than all other age categories; a lower proportion of these patients reported understanding the information to be ‘very easy’ (40%) compared to all other age categories (ranged between 60-67%). However, the number of patients aged 80 years and over was small and when ‘very easy’ and ‘easy’ responses were combined there was little difference across age categories. Patient ratings of helpfulness of the information was also statistically significantly lower for older age categories (p=0.005), with similar patterns of responses to understanding the information. Patients from minority ethnic groups reported less ease in understanding information provided; 81% of patients from a minority ethnic group reported understanding the information to be ‘easy’ or ‘very easy’ compared to 96% of patients from white ethnic groups.

The quantitative survey analysis did not indicate any other clear differences in accessibility of patient information across groups, however qualitative interviews identified other factors affecting the accessibility of patient information. Several patients noted it was particularly difficult to take on the information when they were unwell, while those with visual impairments would have welcomed written information in a larger font. In some cases, patients reported they had received little information about the service or would have liked additional information about the service, their condition and recovery (e.g. information relating to the time of day to take readings, how to interpret oxygen saturation readings, how to manage their recovery, and where to seek further help).

#### Patients’ ability to complete tasks as part of the service

When enrolled on the service, patients were required to complete monitoring activities (i.e. using an oximeter, and recording and providing readings) and escalation activities (i.e. seeking further help). We found evidence of differences in patient reported ease of completing tasks with age, gender, health status, ethnicity, and level of education (see Appendix 7).

#### Monitoring activities

Survey analysis found evidence of a difference in patient-reported ease of recording readings depending on whether they were limited by a pre-existing health problem/disability or not (p=0.004). Fewer patients with a health problem or disability rated recording readings as ‘very easy’ (70%) compared to those without a health problem (79%), however when this response option was combined with ‘easy’ there was little difference between groups. A similar trend was found for health status and providing readings to the service (p=0.017).

There was evidence of differences between patients from white and minority ethnic groups for recording readings (p<0.001). Fewer patients from minority ethnic groups reported recording readings (91%) to be ‘easy’ or ‘very easy’ compared to white ethnic groups (98%). A similar trend was found for providing readings to the service (p=0.013); fewer patients from minority ethnic groups reported providing readings (91%) to be ‘easy’ or ‘very easy’ compared to white ethnic groups (98%). Communication challenges between patients and staff might be one reason for the differences in ease of engagement across ethnic groups (as shown by minority ethnic groups reporting more difficulty understanding the service information). Staff interviews indicated translation services were often needed and not always available for patients for whom English was not their first language. Services often relied on liaising with friends or family to support communication (when translation services were not available) and this was not always possible.

The survey analysis did not find any statistically significant differences in monitoring activities across age categories (at the p<.01 level), however there was some evidence of differences in ease of recording (p=0.022) and providing readings (p=0.016) with age. There was little difference in the proportion of patients reporting tasks as ‘easy’ or ‘very easy’ across age categories, however fewer patients aged over 80 years reported recording readings (55%) and providing readings (57%) to be ‘very easy’ compared to those under 50 years (80%, 79%) and in the 50-64 years age category (77%, 78%). Patients over 80 years of age also tended to report support from friends, family and healthcare professionals was more important for their engagement with the service than younger age groups. Twenty-six percent (n=11/43) of patients over 80 years reported they had support from family and friends to use equipment and 49% (n=21/43) reported support from healthcare professionals helped them to engage with the service. This compares to 21% (n=40/195) of patients under 50 years of age reporting support from family and friends, and 43% (n=83/195) reporting support from healthcare professionals helped them to engage with the service.

A statistically significant difference in providing readings to the service (p=0.001) was also identified for gender. While similar numbers of males and females reported providing readings to the service as ‘easy’ or ‘very easy’ (when responses combined), fewer male patients reported providing readings (70%) to be ‘very easy’ compared to female patients (80%). A higher proportion of males (25%; n=97/385) also reported the importance of friends and family in supporting their engagement with the service compared to females (19%; n=102/531).

There were no statistically significant differences (at the p<.01 level) in ease of engagement with monitoring activities depending on whether patients were employed or not. In qualitative interviews several patients who were in paid employment reported that they would prefer to reduce the number of readings they had to give each day or that they preferred to receive calls at a specific time each day. Similarly, survey analysis did not find statistical evidence of a difference in patient rated ease of monitoring activities with living situation (at the p<.01 level). However, patients living with others tended to report greater reliance on support networks, friends and family. Ten percent (n=14/137) of patients living alone reported support from family or friends encouraged/helped them to engage with the service, while 23% (n=175/754) of those living with others reported support from others to be important. Feedback from patients found that many reported the support from family members, friends or carers to take and provide readings, and liaise with the remote monitoring team was crucial, particularly for patients who were more acutely unwell.

Overall, survey analysis found that most patients reported monitoring activities ‘easy’ or ‘very easy’ to engage with – the proportion reporting difficulties was low. However, findings also indicate that some patient groups might require additional support to engage with the service. This is consistent with patient qualitative data. Some patients reported receiving additional support beyond the monitoring protocol or national guidance (e.g. face-to-face visits). The extent of support needed might be related to severity of illness; several patients reported they were too unwell to use the oximeter themselves or submit readings, requiring additional support from others.

#### Escalation processes

As for escalation processes, survey analysis found evidence of a difference with employment status (p=0.007). Fewer patients who were not in employment reported seeking further help to be ‘easy’ or ‘very easy’ (84%) compared to those who were employed (89%). There was also some evidence of differences in ease of engagement with escalation processes across ethnic groups (p=0.015) and with level of education (p=0.015), however these were not statistically significant (at the p<.01 level). A smaller proportion of patients form minority ethnic groups (76%) reported seeking further help to be ‘easy’ or ‘very easy’ than patients from white ethnic groups (88%), and a smaller proportion of patients (83%) educated to A level, degree level or equivalent reported seeking help to be ‘easy’ or ‘very easy’ compared to patients with a lower level of educational attainment or no qualifications (91% of patients educated to GCSE level or equivalent and 94% with no formal qualification).

### Disparities in patient engagement by the mode of service

Qualitative data found most patients were satisfied with processes for submitting readings (whether tech-enabled or analogue). Patients reported in interviews that they were often not given a choice about the mode of monitoring, but most were happy with the option given and patients that did not have digital infrastructure and/or skills valued the analogue option.

Overall, 25% (n=228/936) of patients reported at least one problem completing tasks that were part of the service (e.g. using the oximeter, providing or recording readings, or seeking further help). Logistic regression analysis (with whether patients reported a problem as the dependent variable) found mode of monitoring was not related to whether patients reported a problem. Nor were health status or ethnicity. Age and education were associated with likelihood of reporting a problem (see Table 4); increasing age and a higher level of educational attainment were associated with an increased likelihood of reporting a problem. The odds of reporting at least one problem with the service were approximately 7.6 times higher for of older adults (over 80 years of age) than younger adults (under 50 years of age), for patients aged 65-80 years the odds were 2.9 times higher and for patients aged 50-64 years were 2.3 times higher compared to patients under 50 years. The odds of patients who were educated to AS, A level, degree level or equivalent reporting at least one problem were 3 times higher than patients who had received no formal qualification.

**Table 4.** Multivariable logistic regression for patient reported problems to the service

| Variable | Participants | Participants reporting a problem (%) | Odds ratio | 95% CI (Lower-upper) | P-value |
| --- | --- | --- | --- | --- | --- |
| Health status |  |  |  |  | 0.727 |
| Not limited at all | 482 | 114 (23.7) | Ref. |  |  |
| Limited a little or a lot | 345 | 87 (25.2) | 1.072 | 0.725-1.585 |  |
| Age |  |  |  |  | <0.001 |
| Younger than 50 years | 195 | 40 (20.5) | Ref. |  |  |
| 50-64 years | 428 | 95 (22.2) | 2.295 | 1.314-4.009 | 0.003 |
| 65-79 years | 256 | 71 (27.7) | 2.910 | 1.606-5.273 | <0.001 |
| 80 years and over | 43 | 15 (34.9) | 7.639 | 2.869-20.339 | <0.001 |
| Education |  |  |  |  | <0.001 |
| No formal qualifications | 146 | 26 (17.8) | Ref. |  |  |
| GCSE level or equivalent | 267 | 46 (17.2) | 1.338 | 0.728-2.460 | 0.348 |
| AS level, A level, degree or equivalent | 319 | 102 (32.0) | 3.039 | 1.726-5.351 | <0.001 |
| Ethnicity |  |  |  |  | 0.081 |
| White ethnic group | 830 | 195 (23.5) | Ref. |  |  |
| Minority ethnic group | 74 | 23 (31.1) | 1.794 | 0.931-3.457 |  |
| Mode of monitoring |  |  |  |  | 0.123 |
| Analogue | 435 | 96 (22.1) | Ref. |  |  |
| Tech-enabled | 501 | 132 (26.3) | 1.363 | 0.920-2.021 |  |

### Disparities in patients’ reports of their experience of the service

Patients reported a positive experience of the service; 93% rated the service as good or excellent, 90% as helpful and 91% reassuring. Survey analysis found evidence of a difference in patient reported experience and helpfulness of the service with age and employment status, however reassurance provided did not differ markedly across any patient groups.

There was evidence that older patients reported the service to be less helpful (p=0.004); fewer patients aged 80 years and over reported the service to be helpful or very helpful (88%) compared to those in the under 50-years (92%) and 50-64 years age categories (92%). Older patients also reported a less positive experience of the service (p=0.034), however this was not statistically significant (at the p<.01 level). The less positive experiences of patients aged 80 years and over might well be explained by differences in understanding information about the service and ease of completing the monitoring tasks, however they might also be due to the smaller sample size of the over 80 years age category.

Patients who were employed rated the service more positively (p=0.001) and found the service significantly more helpful (p=0.003). A higher proportion of patients in employment (96%) rated the service to be good or excellent than those not in employment (92%), and more patients rated the service as helpful or very helpful (93%) compared to those not in employment (87%). The differences in patient experiences based on employment status might be related to other factors, such as health status, level of education or age, with older patients more likely to be retired.

Living situation was not identified in the survey data to be a significant factor in patient reported experience (p=0.912) or reassurance provided (p=0.484). However patient interview data indicated living situation and strength of social network as important factors; patients reported the reassurance provided by the service to be particularly important for those socially isolated or living alone.

## Discussion

### Summary of findings

Our findings indicate at a local level many sites designed and adapted their service to be more inclusive to the needs of their local population, to expand its reach and facilitate engagement. Strategies included broadening the service inclusion criteria to include at-risk groups, setting up referral pathways across services, providing information in different languages or formats, offering translation services, offering analogue and tech-enabled options for relaying readings, face-to-face assessments, delivering oximeters, providing additional training or support, flexible monitoring processes, and liaising with family or friends. However, there was considerable local variation in the adaptations employed by services to meet the needs of specific patient groups. The adoption of such strategies was often dependent on local leaders as to how patients were approached and supported^32^, and often dependent on resources/capacity.

Despite efforts to adapt services, our evaluation found disparities across patient groups in their experience of, and engagement with, the service. Age and employment status were statistically significant factors in explaining patient experience. Age and ethnicity were associated with patient reports of understanding information. Age and level of education were related to whether patients reported a problem with the service, and health status, ethnicity, gender, and level of education were associated with engagement with monitoring and escalation.

### How findings relate to previous research

Despite previous evaluations investigating models of remote home monitoring, and implementation of such services for COVID-19 patients, to our knowledge, this is the first research that has focused on the strategies adopted by services to address disparities in patients’ access, engagement and experience. Our findings were consistent with previous research that highlighted a variety of strategies can be used to reduce health disparities^18,20,23^. For example: targeting resources to at-risk groups by expanding the admission criteria and referral routes, and ensuring digital inclusion by offering analogue options of monitoring. The availability of an analogue option is important given the overlap between patients at greater risk of severe illness from COVID-19 and of digital exclusion. Our finding that mode of service did not have an impact on patient engagement might be explained by the fact that all sites offered analogue modes, or that patients were generally satisfied with the mode offered; indicating staff were able to appropriately assess patients’ digital literacy and infrastructure.

Our findings extend research by outlining some strategies not previously identified e.g. staff providing additional support beyond monitoring (linking with social support or more contact time), amending protocols to patient need and liaising with family/carers. Some possible areas for improvement included the need to: routinely collect and analyse data on relevant populations to check representativeness of uptake, provide accessible information, co-develop platforms with patients and staff, and work with communities to promote the service. Findings support research that has shown similar trends in patients’ experiences of accessing and using health services more generally - relating to ethnicity, deprivation and age and considering differences in expectations, health literacy, digital skills, staff-patient communication, and understanding how systems work^33–35^.

Our findings build on assumptions of the NIA framework^26^ that patient engagement with, and experiences of remote monitoring services differ with fundamental, socio-economic and geographic factors, and support the assumptions that addressing health disparities in designing and delivering services requires a multidimensional perspective. The identification of these factors as related to healthcare experiences can help to understand the mechanisms that create and sustain disparities, shape the design and delivery of remote models of care and guide evaluations monitoring the impact of services on health equity.

### Implications and recommendations

Our evaluation generates a range of recommendations for the design and delivery of remote monitoring services not only for COVID-19 patients, but also other health conditions. Services must consider the needs of their local population and adapt services accordingly, monitor the impact of the service on population groups at risk of health disparities, and work with existing local systems (e.g. community groups and local authority links) to engage harder to reach groups.

Given that many of the groups of patients (e.g. clinically vulnerable, older adults, those without support or lower health literacy) that services aimed to reach found it more difficult to engage with the service, patients’ needs and circumstances should be assessed upon referral in a standardised way and services must be tailored to provide appropriate levels of support for patients and carers. Examples of appropriate tailoring include: patient information in a range of formats to increase accessibility^36^, additional information, education or support to use equipment or in navigating services, flexible and personalised monitoring processes, a non-digital option for patients without the relevant digital skills or infrastructure. Previous research has shown appropriate tailoring of health interventions can promote health equity and flexibility is key for responding to individual needs^37^. Where relevant, services should encourage the involvement of family, friends or carers, however should provide them with the adequate tools and support to reduce the burden of care placed upon them^38^.

When implementing remote models of care, adequate resources and infrastructure should be provided to allow patients to be involved in the design and implementation. Co-design can help to ensure services are better suited to patient needs, and facilitate adherence and engagement^39^. Holistic healthcare and collaboration or linking between services can help to reduce inequalities, particularly for patients who are unwell or have comorbidities; previous research found linking between services promotes flexibility to respond to patient needs^37,39^.

### Strengths and limitations

Our research has several strengths which add to the validity of our findings. The mixed methods design allowed for the triangulation of data across data sources. The evaluation was conducted across several sites and the large number of staff and patients sampled increase the generalisability of findings. The evaluation team was multidisciplinary, which facilitated triangulation and interpretation.

However, limitations should be noted. Several patient groups were underrepresented in the survey when compared to national onboarding data (see results section). In addition, the response rate for the patient survey was relatively low (17.5%) so some impact of selection bias cannot be ruled out. Findings might not be representative of all patient groups and experiences; such as those not referred, who declined or disengaged from the service. The analysis of experience and engagement across patient groups could have been subject to false positives due to the number of comparisons made, although a more stringent p-value was used (p<.01) in reporting significant results. When making comparisons between patient groups, the sample size of several groups was relatively small (e.g. patients aged over 80 years). Qualitative interviews with patients focused on experiences of the service more broadly and not necessarily inequalities, so much of our analysis draws on staff perspectives or patient survey data.

### Future research

There is little published literature on the implementation of remote monitoring and health disparities. There are other populations at risk of health disparities that were beyond the scope of this evaluation and should be considered in any future evaluations – for example, the homeless community, people with disabilities, people with a mental health diagnosis, or people with a visual or hearing impairment. The scope of our evaluation did not allow us to examine the impact of specific strategies made by local services to increase accessibility or engagement and therefore we are unable to make any conclusions about their effectiveness for particular patient groups. Nor are we able to determine the workforce or resource implications of specific strategies. This is an area for future research.

### Conclusion

When health services undergo such a rapid transformation, as we have seen with the shift towards remote models of care during the COVID-19 pandemic, evaluations of the effectiveness of such services must include an impact assessment to ensure the service is accessible and inclusive to all patients and to monitor the impact on health disparities. Addressing health disparities must therefore be a key focus in the planning, design and delivery of remote monitoring models for COVID-19 and other conditions, both nationally and locally, so that remote models of care can be of value to all population groups. National guidance should support services to be inclusive and flexible to best serve the needs of their local population. Staff and patients from groups typically experiencing disparities in their healthcare must play an active role in service design to ensure their needs, experiences and expectations are accounted for.

## Data Availability

Due to the consent process for data collection within this evaluation, there are no data that can be shared.

## Declarations

## Acknowledgements

We are indebted to all of the services who participated in this study and to all of the patients, carers and staff who participated in our surveys and interviews. Thank you to the following: Dr Jennifer Bousfield for supporting with study design and data collection, Simon Barnes for supporting with data entry; Steve Morris and Jon Sussex for advice given throughout the project; our NIHR BRACE and NIHR RSET public patient involvement members; the NIHR 70@70 Senior Nurse research Leaders for providing feedback on the development of our study; Russell Mannion for peer-reviewing our study protocol; Cono Ariti (Cardiff University) for statistical advice; and the NIHR Clinical Research Networks for supporting study set up and data collection.

We thank the NHS Digital CO@h Evaluation Workstream Group chaired by Professor Jonathan Benger and NHSX (in particular Breid O’Brien and Chris Richmond) for facilitating and supporting the evaluation, and to the other two evaluation teams for their collaboration throughout this evaluation: i) Institute of Global Health Innovation, NIHR Patient Safety Translational Research centre, Imperial College London and ii) the Improvement Analytics Unit (Partnership between the Health Foundation and NHSEI).

Many thanks to our Clinical Advisory Group for providing insights and feedback throughout the project, Dr Karen Kirkham (whose previous role was the Integrated Care System Clinical Lead, NHSE/I Senior Medical Advisor Primary Care Transformation, Senior Medical Advisor to the Primary Care Provider Transformation team), Dr Matt Inada-Kim (Clinical Lead Deterioration & National Specialist Advisor Sepsis, National Clinical Lead - Deterioration & Specialist Advisor Deterioration, NHS England & Improvement) and Dr Allison Streetly (Senior Public Health Advisor, Deputy National Lead, Healthcare Public Health, Medical Directorate NHS England).

## Contributors’ statement

All authors were responsible for the study conception, design, and data collection throughout the study. MS and LH led qualitative data analysis and NC led survey analysis. NC, LH and MS drafted the manuscript with contribution from all authors. All authors commented on drafts of the manuscript and approved the final version. NJF was principal investigator for the study.

## Ethical approval

The staff aspects of the evaluation were categorised as a service evaluation by the HRA decision tool and UCL/UCLH Joint Research Office and received ethical approval from the University of Birmingham Humanities and Social Sciences ethics committee (ERN_13-1085AP39). The patient aspects were reviewed and given favourable opinion by the London-Bloomsbury Research ethics committee (REC reference: 21/HRA/0155).

