## Supplementary material for "Examining disparities relating to service reach and patient engagement with COVID-19 remote home monitoring services in England: a mixed methods rapid evaluation"

### Appendix 1. Methods for rapid evaluation of patient and staff experiences of COVID-19 remote home monitoring

#### NATIONAL SURVEYS OF IMPLEMENTATION AND PATIENT AND STAFF EXPERIENCES

The aim of the national study of implementation and patient and staff experiences was to a) understand the development of COVID-19 remote home monitoring services and b) analyse the implementation of COVID-19 remote home monitoring services, and patient and staff experiences of care in sites across England.

##### Sample and recruitment

###### *Selection of sites*

Twenty-eight services were included in our national evaluation. Each site had a research lead (MS, CVP, HW, JB, IL, LH, NC) to support data collection and act as an ongoing point of contact for the site.

To obtain maximum variation, we sampled services based on a range of criteria, including the setting (primary care or secondary care), type of model (pre-hospital, early discharge, both), mechanism for patient monitoring (paper-based, app, both), geographic location (across different areas of the country), timing of implementation (implemented since wave 1 of the pandemic or recently implemented) and involvement in the evaluation with the other evaluation partners (Imperial and IAU).

Sites were recruited through an expression of interest process whereby we presented our study at local and national meetings and asked sites to express interest in participating. Clinical Research Networks facilitated the setup of sites and local governance approvals. Some sites were identified through our phase 1 evaluation.

###### *Staff survey*

We conducted a survey of staff involved in delivering COVID-19 remote home monitoring services in the 28 services, including clinical leads, delivery staff and data staff. Staff at participating sites distributed surveys to staff. All survey sites were asked to keep a record of the number of surveys they have sent out to determine staff response rates. For the staff survey, staff received an email from their GP practice/hospital or other relevant networks (together with reminder emails) with a link to fill out an online survey.

###### *Patient survey*

25/28 sites agreed to conduct the patient and carer survey. To participate in our survey, participants needed to be:

I. 18 or over,
II. Proficient in English (or one of the following languages: Polish, Bengali, Urdu, Punjabi, French and Portuguese),
III. Eligible to receive COVID-19 remote home monitoring services, and must also have been offered and received COVID-19 remote home monitoring.

National and local eligibility for COVID-19 remote home monitoring varies. We were flexible within our sampling to take into account both national and local eligibility criteria. For reference, the national eligibility guidelines are as follows: To be eligible for receiving COVID-19 remote home monitoring patients must have a confirmed or suspected diagnosis of COVID-19 plus be one of the following: (a) Symptomatic with COVID-19 & aged 65 years or older, b) Symptomatic with COVID-19 & under 65 years but 'clinically extremely vulnerable' (using the Clinically extremely vulnerable to COVID list) to COVID (CO@h Standard operating procedure, 2020).

For the patient survey, NHS staff from participating services sent the patient survey to patients (or their carers if applicable) onboarded onto the service between 1st January 2021-11th June 2021. NHS sites decided how to best disseminate the survey to their patients (either via post or text/email). All survey sites were asked to keep a record of the number of surveys they have sent out to determine patient response rates.

If patients were not able/willing to take part in the survey, they were given the option to ask their carer or family member to complete the survey on their behalf, reflecting on the patient's experience with the service. The survey was sent to patients who have received care at participating sites by NHS staff. Patients/carers returned completed surveys directly to the study team for analysis, either electronically through REDCap or via post using pre-paid envelopes. In addition to English, we also offered participants the opportunity to receive an information sheet and survey in six other languages (Polish, Bengali, Urdu, Punjabi, French and Portuguese).

#### Measures

##### *Staff survey*

We developed staff surveys specifically for this study. Different sets of questions were developed for different groups of staff (i.e. one survey for service leads and one survey for staff delivering the service). The main purpose of the staff survey was to gather information on the staff involved in delivering COVID-19 remote home monitoring services, different set-up processes and models implemented, staff experiences of implementing these models, factors influencing delivery and staff perceptions of patient engagement with the service. As part of the survey, we also sought to explore different experiences of analogue vs tech-enabled models. The survey included a number of closed questions which focused on documenting staff experiences of setting up, managing and delivering the service. These questions were followed by a single, open text question at the end to give staff the opportunity to share any wider thoughts. To reduce burden and maximise response rates, the online survey was developed to take no longer than 15-20 minutes to complete. The survey was delivered using an online platform (REDCap).

Survey questions were reviewed, and sense-checked by our clinical advisory group, PPI group and our cohort of 70@70 nurses (senior nurse and midwife clinical leaders with demonstrable experience of building a research-led care environment for patients). The theoretical frameworks were used as a sensitising device to inform the development of questions in the surveys and interviews. The staff survey was piloted with a small number of sites. Piloting aimed to determine whether questions were appropriate and relevant, while identifying areas for further refinement prior to circulation nationally. In response to feedback, we amended some of the staff survey questions and response option wording to improve clarity, added response options, re-ordered questions and amended question format.

##### *Patient survey*

We developed patient surveys specifically for this study. The aim of the patient survey was to capture the experiences of patients who received COVID-19 remote home monitoring, and their engagement with the COVID-19 remote home monitoring service. We developed a patient experience survey for this purpose. The survey included closed questions focused on: the service that patients have received, their experience with the service and their engagement with the service. As part of the survey, we also asked questions about patients experience of analogue vs tech-enabled models. These questions were followed by a single open text question at the end to give participants the opportunity to share any wider thoughts. Survey and interview questions were informed by relevant service documentation (NHS, 2020, NHS 2021), theoretical frameworks relating to social, political and technical contexts (Greenhalgh, 2017; Powell, 2010; Greenhalgh, 2015; Lehoux & Blume, 2000) and behaviour (Michie et al, 2011), and previous literature on engagement [Borrelli, 2011; Walton et al, 2017]. We also included a section at the end of the survey to ask about participants' socio-demographic characteristics (including questions on gender, age, ethnicity, education, employment, disability, sexuality, first language and geographical region). Questions on demographic characteristics were informed by previous literature (Lyratzopoulos, 2012; Pianori, 2020; ELSA, 2019; Wenz 2021; ONS, UK government, 2018). To reduce burden and maximise response rates, the patient survey was designed to take between 15 and 30 minutes to complete. The survey was delivered using the online platform REDCap.

Survey questions were reviewed, and sense-checked by our clinical advisory group, PPI group and our cohort of 70@70 nurses. The theoretical frameworks were used as a sensitising device to inform the development of questions in the surveys and interviews. The patient survey was piloted with our PPI group and some members of the public. In response to feedback for the patient survey, and to make the survey more accessible, we amended some of the questions, increased the font size, reduced the number of questions and added definitions for key terms (such as oximeter).

#### Data collection

##### *Staff survey*

For the staff survey, staff were asked to follow an online link to complete the survey. If they followed the link, they reached an information page which provided background to the study, potential risks and a description of how the data will be used to ensure informed and voluntary participation. It was emphasised that individual responses would be treated confidentially and reported anonymously. Staff were asked to tick a box to indicate their consent to take part in the study. Data collection took place between February and May 2021.

##### *Patient survey*

For the patient survey, patients were approached by NHS staff to take part in a survey in one of two different ways: 1) if the patient was monitored through the use of an app, they received an SMS/email with a link to the online survey, 2) if the patient was monitored through regular phone calls and a paper-based recording method, they received the survey in the post (with a pre-paid addressed envelope). Whilst most surveys were distributed at discharge, some sites chose to distribute the paper survey at onboarding and then remind patients at discharge to complete the survey. NHS staff distributed the online and paper version of the survey so the research team had no access to patient information. Both survey options (online and paper) included prefacing information with a background to the study, potential risks, indicating voluntary participation, anonymity and a description of how the data will be used. This page also included boxes that patients/carers were asked to tick to indicate their consent to take part in the study. The method of administering the survey to patients (i.e. NHS staff sending to patients) meant that there were no reminders. Due to research capacity, we were unable to conduct the survey with patients over the phone. Data collection took place between March and June 2021.

#### Data management

Surveys were returned to the research team, either electronically through REDCap (staff and patient surveys), or by posting completed surveys in pre-paid envelopes to our RSET team members at the Nuffield Trust or UCL (patient surveys only). Surveys received via post were stored securely in locked filing cabinets within secure Nuffield Trust or UCL offices. Data from patient surveys sent via post were inputted into REDCap by members of the research team. Data from the patient surveys were directly stored in the UCL Data Safe Haven via REDCap, as this will include identifiable information (e.g. postcode data). Data from the completed surveys were stored securely using password protected spreadsheets to which only the RSET and BRACE researchers had access to.

#### Analysis

We analysed staff survey data using SPSS statistical software (version 25). Descriptive statistics including frequencies and percentages were calculated to explore staff perceptions of patient groups facing barriers. Service lead responses (one selected from each site based on their role) were analysed to determine the number of sites that made service adaptations according to patient needs or requirements. Open text responses relating to local service adaptations and patient groups facing barriers were analysed; responses were triangulated with service lead interview data, coded and grouped into themes related to service design, delivery and adaptations.

For patient survey data, we used descriptive statistics including frequencies and percentages. Analysis was organised according to three factors of the NIA health disparities framework; fundamental, socioeconomic and geographic. Non-parametric univariate analyses were conducted to explore patient engagement with the service (e.g. accessibility of the information, achievability of tasks forming the service, and problems completing tasks) across the fundamental factors; age, gender, ethnicity, and health status, socio-economic factors; education, employment status, English as first language, and living situation, and the geographic factor; deprivation score. We selected non-parametric tests due to the skewed nature of the survey data and the fact that dependent variables were Likert-type scales consisting of ordinal data. We used Mann-Whitney U tests when independent variables comprised only two levels and Kruskal-Wallis H tests for more than two levels when the dependent variable comprised ordinal data. Due to the large number of statistical hypothesis tests conducted, and hence the possibility of false positive results at the traditional level of statistical significance of $p<0.05$, it was determined that a statistical significance level of $p<0.01$ should be used. If statistical significance was detected across more than two groups, post hoc analyses were conducted. Similar non-parametric univariate analyses were used to explore patient views and experiences of the service across the same fundamental, socio-economic and geographic factors.

We conducted logistic regression modelling to examine whether modality of the service (i.e. whether tech-enabled or analogue), age, education, health status and ethnicity were associated with likelihood of patients reporting at least one problem with the service. We reported odds ratios (ORs) with 95% confidence intervals for the regression models. Patient open text survey responses providing feedback about the service and any suggested changes or improvements were triangulated with patient interview data, coded and grouped into themes related to service design and barriers to engagement.

#### CASE STUDIES OF IMPLEMENTATION, STAFF AND PATIENT EXPERIENCES

The aim was to document the implementation of COVID-19 remote home monitoring services (including the identification of factors acting as barriers and enablers in implementation), staff experiences of delivering the service and in-depth patient experiences of care in a sample of 17 sites. All 17 sites also took part in the staff survey and 15/17 sites took part in the patient survey.

##### Sample and recruitment

*Site selection*

A smaller sample of the overall study sites were included as case studies in order to conduct a more in-depth analysis of implementation, patient and staff experiences. Seventeen of the twenty-eight sites were selected as in-depth case study sites using the aforementioned criteria (see national site selection). Four of the 17 sites were purposively selected by NHSX for a more in-depth analysis of implementation and patient and staff experiences of tech-enabled models of care; sites using different tech-enabled platform were selected.

*Staff interviews*

For staff interviews, we aimed to purposively sample one or two members of staff delivering the service, one staff member leading the service (operationally or clinically), and one staff member knowledgeable about service data collection/analysis at each of the 17 sites (note, one staff member may fulfil more than one role).

Participants for the staff interviews were approached through each case study site's contact person/gatekeeper. Potential interviewees were introduced to the researcher or asked to contact the researcher to take part. The researcher contacted these potential participants via email and sent them a participant information sheet. Participants were given 48 hours to review the information and ask questions about the study. If the participant agreed to take part in the study, they were asked to sign the consent form. An informed consent process using participant information sheets and written consent (scanned forms or typewritten/electronic signature) was used for recruitment to ensure informed and voluntary participation.

##### *Patient interviews*

We aimed to interview up to six participants (patients or their carer) who had received, disengaged or declined with COVID-19 remote home monitoring from each site. To participate in our patient or carer interviews, participants needed to be 18 or over, proficient in English (or one of the following languages: Polish, Bengali, Urdu, Punjabi, French and Portuguese), eligible to receive COVID-19 remote home monitoring services, and must also have been offered and either received or refused the service. If patients were not able/willing to take part in the interview, patients were asked by site coordinators if their carer (if they have one) could be approached to capture their perceptions of the patient's journey and overall experience with the service.

We asked the main contact person at each site (the study coordinator), to identify a convenience sample of 4-6 patients (or their carers). To identify potential participants, the study coordinator contacted potential participants to see if they were happy to be approached by a researcher. If they agreed, the researcher contacted the patient or their carer via telephone or email to discuss the study. NHS staff identified patients on behalf of the study team using a purposive sampling approach. To be inclusive and capture a wide range of views, we asked sites to select patients with different characteristics (e.g. age, gender, ethnicity, deprivation score (by postcode), employment status, and comorbidities). We also asked sites to identify interviewees who had declined or disengaged from the service and those who used different data submission methods (if applicable).

If the patient or their carer was contacted via phone, they were asked if a participant information sheet and consent form can be sent via email. If they preferred post, both of these documents were sent via post with a pre-paid addressed envelope so they could return the signed consent form to the team. If the patient was contacted via email, the participant information sheet and consent form were sent in a subsequent email and the patient was given the option to schedule a call with the researcher to discuss the study. The participant information sheet contained information on the study, potential risks and a description of how the data will be used to ensure informed and voluntary participation. If the patient or their carer agreed to take part in the study, they were asked to email back the signed consent form (scanned forms or typewritten/electronic signature). If patients were not able/willing to take part in the interview, we asked patients if we can approach their carer (if they have one) to capture their perceptions of the patient's journey and overall experience with the service.

#### Measures

##### *Staff interviews*

Staff interview topic guides included questions for staff leading a service, staff delivering a service and staff involved in data. The staff topic guides included questions about their role, the origin of the model, the aims and goals of the model, resources and processes of the model, staff training, facilitators and barriers of implementation, patient engagement, adaptations, monitoring and evaluation, impact, and recommendations and sustainability.

In the four sites for in-depth analysis of tech-enabled platforms, interviews with delivery staff were extended to include a 'think aloud' section where staff narrate the process of using the platform in situ (think aloud methodology, Eccles & Arsal, 2017).

##### *Patient and carer interviews*

Patient and carer interview topic guides included questions for patients who had received COVID-19 remote home monitoring services, those who had declined to receive the service and those who disengaged from the service. The interviews with patients and carers focused on documenting their journeys of remote home monitoring, their experiences of being ill and monitored at home, experiences with escalation and discharge, their engagement with the service, and recommendations for improving these models. Interview questions (as with survey questions) were informed by relevant service documentation (NHS, 2020, NHS 2021) and literature (Greenhalgh, 2017; Powell, 2010; Greenhalgh, 2015; Lehoux & Blume, 2000; Michie et al, 2011; Borrelli, 2011; Walton et al, 2017).

During the interview, we asked patients/carers some brief questions relating to socio-demographic characteristics including whether they are a patient or carer, age, gender, ethnicity, how many people they live with, education and qualifications, employment status, English as a first language, disability and postcode (the latter to be used as indicator of social deprivation) (Lyratzopoulos, 2012; Pianori, 2020; ELSA, 2019; Wenz 2021; ONS, UK government, 2018). We emphasised that as with all parts of the interview, these questions are optional.

We intended to conduct 'think alouds' with patients who had used tech-enabled data submission from the four sites selected for in-depth analysis of tech-enabled platforms; however, patients did not have access to the platforms after discharge and recall of the use of these platforms during their illness was poor. Consequently, think aloud methodology was discontinued for patient interviews.

To determine whether questions were appropriate and relevant, we discussed the interview topic guides with our PPI members and the 70@70 nurses. The topic guides were amended accordingly.

##### Data collection

The researcher arranged a time to carry out the interview. Each site had a different lead researcher who conducted the interviews and liaised with sites on an on-going basis. Interviews were conducted by six researchers (MS, CV, HW, LH, IL, JB). Interviews were carried out via telephone or an online platform (e.g. Zoom or MS Teams) as preferred by the participant. Data collection for interviews was conducted between February and June 2021.

##### Data management

All interviews were semi-structured, audio recorded (subject to consent being given), transcribed verbatim by a professional transcription service (TP Transcription limited), anonymised and kept in compliance with the General Data Protection Regulation (GDPR) 2018 and Data Protection Act 2018.

##### Analysis

For patient and staff interviews, data collection and analysis was carried out in parallel and facilitated through the use of rapid assessment procedure (RAP) sheets.

To understand whether and how local services were adapted to address health disparities and their potential impacts (RQ1), a thematic analysis was carried out on primary data from service lead interviews. Interview transcripts were read and re-read to become familiar with participants' accounts. Inductive, line-by-line coding was conducted using NVivo 12 software, labelling segments of the data. Each code's data were checked for consistency of interpretation and re-coded as necessary. Codes were organised into higher-order themes related to service adaptations aimed at increasing inclusivity/reach or engagement.

To examine the potential impacts of adaptations from patients' perspectives, rapid assessment procedure (RAP) sheets – summary templates of patients interviews based on questions in the interview guidewere analysed. RAP sheets were read and re-read to become familiar with the data, getting a feel for the range of participants' accounts, recurring experiences, views and problems, and unique experiences. Patient RAP sheets were imported into qualitative analysis software (NVivo12) and data was deductively coded using the framework outlined by the themes from the service lead interviews.

### Appendix 2. Staff & patient survey

#### COVID HOME MONITORING STAFF SURVEY

We would like to invite you to take part in our evaluation of the COVID monitoring at home service (also referred to as the COVID oximetry@home service, COVID care at home or COVID virtual wards).

We are interested to hear the views and experiences of staff involved with the delivery or management of the COVID home monitoring service. Your participation in this survey will provide helpful feedback about being part of the service. The survey will take approximately 5-10 minutes to complete and your feedback will be recorded anonymously.

This national survey is part of a larger piece of work, which aims to explore the impact of COVID care at home, throughout the COVID-19 pandemic. The evaluation is being undertaken by two national Rapid Evaluation Centres: RSET (based at University College London and the Nuffield Trust) and the Birmingham, RAND and Cambridge Evaluation (BRACE) Centre. It is funded by the National Institute for Health Research (NIHR) and NHS England and Improvement.

For further information, please click on the information sheet below.

**By ticking the box below, you are agreeing to take part in this survey:**

I confirm that I have read the information sheet (18/02/2021, version 1.0) for this survey, I understand that my participation in this survey is voluntary, and that data will be anonymised so that I cannot be identified. I consent to taking part in this survey.

What is your involvement in the covid home monitoring service?
- Clinical lead and/or service manager
- Staff member delivering covid home monitoring

##### Clinical lead and/or service manager

***Please answer the below survey questions about covid home monitoring. The first few questions relate to your role within the service.***

1. What is your professional role within the service? (Please select all that apply)
   - Clinical
   - Non-clinical
   - Volunteer
   - Student
   - Other, please specify

2. What is your job or role title?

3. Have you been redeployed from your usual role to work on the covid home monitoring service?
   - Yes
   - No
   - Not applicable

4. Are you sharing your covid home monitoring role with any other roles (such as other clinical responsibilities)?
   - Yes, please specify
   - No
   - Not applicable

5. When was your covid home monitoring service launched?
- Drop down list [March 2020 – April 2021]

6. How long have you been involved in the covid home monitoring service?
- Less than 1 month
- 1-3 months
- 4-6 months
- 7-9 months
- More than 10 months

7. What aspects of the covid home monitoring service have you been involved in? (Please select all that apply)
- Developing and/or piloting
- Set-up and design
- Management
- Referring patients
- Triaging patients
- Patient monitoring
- Escalation
- Discharge
- Monitoring/evaluating the service
- Administration
- Other, please specify
- None of the above

8. For covid home monitoring, where are you based?
- Remotely
- Hot hub
- GP practice
- Hospital
- Other, please specify

***The next questions relate to your covid home monitoring service model***

9. Which setting holds responsibility for the patients within the covid home monitoring service? (Please select all that apply)
- Primary care
- Secondary care / hospital
- Community services
- Other, please specify

10. What type of model is your covid home monitoring service? (Please select all that apply)
- Pre-hospital (referred from community)
- Pre-hospital (referred from Emergency Department)
- Early discharge from hospital
- Other, please specify

11 a. If your covid home monitoring is a pre-hospital (community or ED referral) service, what criteria are used to enrol patients? (Please select all that apply)
- Diagnosed with Covid-19: either clinically or positive test result
- Symptomatic
- Age, please specify
- Clinically vulnerable to Covid (e.g. late presentation, prior respiratory or chronic illness)
- Downs Syndrome

- Other Learning Disability
- Black Asian and minority ethnic
- Obesity
- Immunosuppressed
- Severe mental illness
- Other, please specify
- Not applicable

11 b. If your covid home monitoring service is an early discharge (post-hospital) service, what criteria are used to enrol patients? (Please select all that apply)

- Diagnosed with Covid-19: either clinically or positive test result
- Symptomatic
- Age, please specify
- Clinically vulnerable to Covid (e.g. late presentation, prior respiratory or chronic illness)
- Downs Syndrome
- Other Learning Disability
- Black Asian and minority ethnic
- Obesity
- Immunosuppressed
- Severe mental illness
- Other, please specify
- Not applicable

12. How are patients identified and referred to your covid home monitoring service? (Please select all that apply)

- NHS 111
- Covid Clinical Assessment Service (CCAS)
- Test and Trace
- Hot Hub
- GP
- Emergency Department
- Ambulance service
- Routine testing in care homes
- Hospital inpatient
- Other, please specify

13. How are the patients within the service monitored? (Please select all that apply)

- Paper diary/telephone
- Digital app
- Email
- Other, please specify

14. Is the covid home monitoring service being delivered 24 hours per day, 7 days per week?

- Yes
- No

15. What processes are involved in your covid home monitoring service model? (Please select all that apply)

- Patient triage
- Patient information and training
- Patient monitoring
- Tools for flagging deterioration
- Escalation and referral to other services
- Patient discharge from the ward

- Other, please specify

16. Who is distributing the pulse oximeters for the covid home monitoring service? (Please select all that apply)
    - Clinical staff
    - Non-clinical staff
    - Volunteers
    - Students
    - Other, please specify
    - Not sure

17. Who is monitoring patients as part of the service? (Please select all that apply)
    - Clinical staff
    - Non-clinical staff
    - Volunteers
    - Students
    - Other, please specify
    - Not sure

18. On average, how often are patients contacted to provide readings as part of the covid home monitoring service? (Please provide details for how this differs with symptom severity and service model)

19. Are patients within the covid home monitoring service provided oxygen and/or prescribed medications? (Please select all that apply)
    - Oxygen
    - Dexamethasone
    - Low Molecular Weight Heparin (LMWH)
    - Other, please specify
    - None of the above

20. To what extent do you agree with the following statement: There are enough staff and capacity to deliver the covid home monitoring service as intended.
    - Strongly agree
    - Agree
    - Neither agree nor disagree
    - Disagree
    - Strongly disagree
    - Not sure

21. Which services are patients signposted to during or following discharge from covid home monitoring? (Please select all that apply)
    - GP
    - Community care
    - Other, please specify

22. In your opinion, are there any types or groups of service users facing barriers to accessing the service? (Please select all that apply)
    - Black Asian and minority ethnic
    - Patients with learning disabilities
    - Older adults (65+)
    - Non-English first language
    - Cognitively impaired
    - Informal carers

- Digitally excluded
- Visually or hearing impaired
- Other, please specify
- None of the above

23. Has there been any tailoring of the covid home monitoring service to accommodate specific service user needs or requirements?
    - Yes, please specify
    - No
    - Not sure

***The following questions relate to your experience of delivering the covid home monitoring service including training and support received***

24. When starting your role in covid home monitoring, did you have any relevant experience or did you require any additional training?
    - Yes, I required additional training
    - No, I had relevant experience
    - I had relevant experience but still required additional training

25. Have you received any training or training resources as part of covid home monitoring?
    - Yes
    - No
    - Not sure

26. Are there any additional training resources or training required to deliver the covid home monitoring service?
    - Yes
    - No

27. Have you received support from NHS England and NHS Improvement (or other organisations) regionally or nationally for any of the following? (Please select all that apply)
    - Training for staff
    - Training for patients
    - Setting up and using systems for recording data
    - Guidance for implementation
    - Obtaining pulse oximeters
    - Distributing pulse oximeters
    - Other, please specify
    - Not sure

28. To what extent has your role within the covid home monitoring service impacted on your workload (e.g. whether taking on the role instead of your usual work or in addition to your usual work)?
    - Very negatively
    - Negatively
    - Neutral
    - Positively
    - Very positively
    - Not sure

29. To what extent has your role within the covid home monitoring service impacted on your job satisfaction?
    - Very negatively
    - Negatively

- Neutral
- Positively
- Very positively
- Not sure

30. To what extent has your role within the covid home monitoring service impacted on your work related stress?
- Very negatively
- Negatively
- Neutral
- Positively
- Very positively
- Not sure

***The following questions relate to the impact of the covid home monitoring service and how it is monitored***

31. Do you think the covid home monitoring service is having an impact on the following? (Please select all that apply)
- Reducing patient mortality
- Reducing patient morbidity
- Reducing health inequalities
- Increasing health inequalities
- Early identification of cases of deterioration
- Reducing attendance/reattendance to ED
- Reducing use of ICU/ventilation
- Reducing hospital admissions
- Reducing length of stay in hospital
- Other, please specify
- None of the above

32. What data are you currently collecting or able to review from patients in order to monitor service delivery? (Please select all that apply)
- Pre-existing clinical conditions
- Disabilities
- Usual care needs (e.g. social services)
- Ethnicity
- Oximetry readings
- NEWS scores
- Changes in other clinical conditions
- Hospital admissions or readmissions with COVID or suspected COVID
- 111 calls
- 999 calls
- Mortality
- Patient experiences
- Date of symptom onset
- Date of onboarding
- Source of referral
- Date of offboarding
- Length of stay in the programme or on the virtual ward
- Hospital length of stay
- Other, please specify
- None of the above
- Not sure

33. How helpful have these data been for monitoring progress against your expected outcomes?
- Not at all helpful
- Not very helpful
- Neutral
- Helpful
- Very helpful
- Not applicable

34. Have you used remote home monitoring for service users with other conditions as part of another service?
- Yes, please specify
- No
- Not sure

35. Is there anything else you'd like to tell us about your experience of delivering or managing the covid home monitoring service? (Please write in the box below)

##### Staff member delivering covid home monitoring

***Please answer the below survey questions about covid home monitoring. The first few questions relate to your role within the service***

1. What is your professional role within the service? (Please select all that apply)
- Clinical
- Non-clinical
- Volunteer
- Student
- Other, please specify

2. What is your job or role title?

3. Have you been redeployed from your usual role to work on the covid home monitoring service?
- Yes
- No
- Not applicable

4. Are you sharing your covid home monitoring role with any other roles (such as other clinical responsibilities)?
- Yes, please specify
- No
- Not applicable

5. How long have you been involved in the covid home monitoring service?
- Less than 1 month
- 1-3 months
- 4-6 months
- 7-9 months
- More than 10 months

6. What aspects of the covid home monitoring service have you been involved in? (Please select all that apply)
- Developing and/or piloting

- Set-up and design
- Management
- Referring patients
- Triaging patients
- Patient monitoring
- Escalation
- Discharge
- Monitoring/evaluating the service
- Other, please specify

7. For covid home monitoring, where are you based?
- Remotely
- Hot hub
- GP practice
- Hospital
- Other, please specify

***The next questions relate to your covid home monitoring service model***

8. Which setting holds responsibility for the patients within the covid home monitoring service? (Please select all that apply)
- Primary care
- Secondary care / hospital
- Community services
- Other, please specify

9. What type of model is your covid home monitoring service? (Please select all that apply)
- Pre-hospital (referred from community)
- Pre-hospital (referred from Emergency Department)
- Early discharge from hospital
- Other, please specify

10. In your opinion, are there any types or groups of service users facing barriers to accessing the service? (Please select all that apply)
- Black Asian and minority ethnic
- Patients with learning disabilities
- Older adults (65+)
- Non-English first language
- Cognitively impaired
- Informal carers
- Digitally excluded
- Visually or hearing impaired
- Other, please specify
- None of the above

11. Has there been any tailoring of the covid home monitoring service to accommodate specific service user needs or requirements?
- Yes, please specify
- No
- Not sure

***The following questions relate to your experience of delivering the covid home monitoring service including training and support received***

How have you found delivering covid home monitoring? Please indicate for each statement.

12. Triage processes
    - Very Easy
    - Easy
    - Neutral
    - Difficult
    - Very difficult
    - Not applicable

13. Monitoring patients (e.g. using the app or paper-based system)
    - Very Easy
    - Easy
    - Neutral
    - Difficult
    - Very difficult
    - Not applicable

14. Processes to escalate patients
    - Very Easy
    - Easy
    - Neutral
    - Difficult
    - Very difficult
    - Not applicable

15. The IT systems you are using
    - Very Easy
    - Easy
    - Neutral
    - Difficult
    - Very difficult
    - Not applicable

16. Working with other covid-related services
    - Very Easy
    - Easy
    - Neutral
    - Difficult
    - Very difficult
    - Not applicable

17. Do you feel adequately supported in your role in the covid home monitoring service?
    - Yes
    - No
    - Not sure

18. When starting your role in covid home monitoring, did you have any relevant experience or did you require any additional training?
    - Yes, I required additional training
    - No, I had relevant experience
    - I had relevant experience but still required additional training

19. Have you received training in your area of responsibility as part of the covid home monitoring?

- Yes
- No
- Not sure

20. How confident are you in your ability to carry out your responsibilities as part of covid home monitoring?
- Not at all confident
- Not very confident
- Neutral
- Confident
- Very confident
- Not sure

21. Have you completed the covid home monitoring competency framework?
- Yes
- No
- Not sure

22. Do you feel clear about the requirements of your role?
- Yes
- No
- Not sure

23. Do you feel you have any further training or support needs?
- Yes
- No
- Not sure

24. To what extent has your role within the covid home monitoring service impacted on your workload (e.g. whether taking on the role instead of your usual work or in addition to your usual work)?
- Very negatively
- Negatively
- Neutral
- Positively
- Very positively
- Not applicable

25. To what extent has your role within the covid home monitoring service impacted on your job satisfaction?
- Very negatively
- Negatively
- Neutral
- Positively
- Very positively
- Not applicable

26. To what extent has your role within the covid home monitoring service impacted on your work related stress?
- Very negatively
- Negatively
- Neutral
- Positively
- Very positively

- Not applicable

***The following questions relate to your views of patient engagement and experience of the service***

27. Overall, how have service users engaged with and responded to the covid home monitoring service?
    - Very well
    - Well
    - Neutral
    - Poorly
    - Very poorly
    - Not applicable

***How well do you feel that service users have carried out the following?***

28. Taking measurements using pulse oximeters
    - Very well
    - Well
    - Neutral
    - Poorly
    - Very poorly
    - Not applicable

29. Providing readings over the phone
    - Very well
    - Well
    - Neutral
    - Poorly
    - Very poorly
    - Not applicable

30. Providing readings via the digital app
    - Very well
    - Well
    - Neutral
    - Poorly
    - Very poorly
    - Not applicable

31. Do you think that service users have felt reassured by receiving covid home monitoring?
    - Very reassured
    - Reassured
    - Neutral
    - Not very reassured
    - Not at all reassured
    - Not applicable

32. Do you think the covid home monitoring service is having an impact on the following? (Please select all that apply)
    - Reducing patient mortality
    - Reducing patient morbidity
    - Reducing health inequalities
    - Increasing health inequalities

- Early identification of cases of deterioration
- Reducing attendance/reattendance to ED
- Reducing use of ICU/ventilation
- Reducing hospital admissions
- Reducing length of stay in hospital
- Other, please specify
- None of the above

33. Is there anything else you'd like to tell us about your experience of delivering or managing the covid home monitoring service? (Please write in the box below)

#### COVID CARE AT HOME: PATIENT AND CARER EXPERIENCE SURVEY

**Background information**

Please read the information sheet and this information before completing the survey.

Thank you for taking the time to look at our survey. The responses you provide will help us to understand more about the experiences of patients and carers (such as family members or friends who have provided care) who have received COVID care at home. The survey is part of a larger national study which is looking at the impact of COVID care at home.

The survey may take between 15 and 30 minutes to complete. The questions in the survey will ask you about the service that you (or your family member) received and your experience receiving and engaging with the service, and some questions about you (and your family member - if applicable). Please feel free to complete this survey with your family member/friend if you would like to. Please do not include any identifiable information (e.g. your name) in your responses.

**After completing the survey, please post your survey responses back to us (the research team) using the pre-paid envelope provided**.

Thank you so much for taking time to read this information. If you would like to complete this survey, please tick all of the statements on Page 2 ('Section A. Consent to take part') and continue to complete the survey.

##### Section A. Consent to take part

By ticking each box below and returning the survey to the research team, you are providing consent to take part in this survey. **Please tick all boxes if you would like to take part** (1-6):

| | Please tick: |
|---|---|
| 1. I have read the information provided for this study (information sheet dated 10/02/2021, version 1.2). I have had the opportunity to consider the information, ask questions and have had these answered satisfactorily. | ☐ |
| 2. I understand that my participation in this survey is voluntary and that I am free to withdraw up to 14 days after completing the survey. I understand if I withdraw, the data I have provided up until that time will be deleted. | ☐ |
| 3. I understand that all personal information will be used for purposes explained to me, will remain confidential and that all efforts will be made to ensure I cannot be identified. | ☐ |
| 4. I understand that any identifiable data will be stored securely and will not be shared with anyone outside of the research team. | ☐ |
| 5. I give permission for the identifiable data I provide to be archived at University College London for up to three years after the end of the project. The anonymised data will be archived for up to 20 years. | ☐ |
| 6. I agree to take part in this survey. | ☐ |

If you have ticked boxes 1-6 above, please proceed to Section B.

#### Section B. Type of respondent

**1.** Are you the patient who received COVID care at home or a carer (such as a family member or friend?) *(please tick)*

☐ Patient or service user

☐ Carer (e.g. family member or friend of someone who has received COVID care at home)

#### Section C. Your experience of COVID care at home

*Please answer the below survey questions about your experience of COVID care at home. The first questions relate to the service you received from COVID care at home.*

**Note for carers completing this survey: If you are answering this survey as a carer, please respond to the questions in this section on behalf of your family member/friend who received COVID care at home (i.e. 'you' refers to you and your family member/friend).*

**2.** How were you referred to the COVID care at home service? *(please tick)*

- ☐ By a community healthcare professional (e.g. GP or community clinics)
- ☐ By the hospital (after attending the emergency department)
- ☐ By the hospital (after being discharged as an inpatient)
- ☐ Other (please specify): ____________
- ☐ Not sure

| | **Yes** | **No** | **Not sure** |
|---|---|---|---|
| **3.** Did you receive an oximeter? (please tick) *(*An oximeter is a small machine that is placed on your finger. Oximeters are used to measure your blood oxygen levels)* | ☐ | ☐ | ☐ |
| **4.** Were you given information on how to use the oximeter? *(please tick)* | ☐ | ☐ | ☐ |
| **5.** Were you given information on how to take and record your readings? (please tick) *(*Readings include oxygen saturation levels, temperature or heart rate)* | ☐ | ☐ | ☐ |
| **6.** Were you given information about what to do if your oxygen levels dropped below the recommended levels? *(please tick)* | ☐ | ☐ | ☐ |

**7.** How was the information provided to you? *(please tick)*

- ☐ Written information
- ☐ Over the telephone
- ☐ Online information (e.g. video)
- ☐ Other (please specify): ____________
- ☐ Not applicable

**8.** How did you find understanding the information about COVID Care at home that you were provided with? *(please circle one response)*

| | | | | | |
|---|---|---|---|---|---|
| Very easy | Easy | Neutral | Difficult | Very difficult | Not applicable |

**9.** How helpful did you find the information on how to use the oximeter? *(please circle one response)*

| | | | | | |
|---|---|---|---|---|---|
| Very helpful | Helpful | Neutral | Not very helpful | Not at all helpful | Not applicable |

| | **Yes** | **No** | **Not applicable** |
|---|---|---|---|
| **10.** Did you have anyone (such as a family member or friend) who could help you to use the oximeter if needed? *(please tick)* | ☐ | ☐ | ☐ |

| | **Paper diary** | **Digital app** |
|---|---|---|
| **11.** How were you asked to record your symptoms and outcomes *(please select all that apply)* | ☐ | ☐ |

| | **Yes** | **No** |
|---|---|---|
| **12.** Were you given the choice between using a digital app or a paper diary? *(please tick)* | ☐ | ☐ |
| **13.** Did you use the same method of recording your symptoms and outcomes throughout? *(please tick)* | ☐ | ☐ |

| | **Yes** | **No** | **Not applicable** |
|---|---|---|---|
| **14.** Were you happy to record using a paper diary? *(please tick. If you didn't use a paper diary, please choose not applicable)* | ☐ | ☐ | ☐ |

**15.** How did you find using the paper diary? *(please circle one response. If you didn't use a paper diary, please choose not applicable)*

| | | | | | |
|---|---|---|---|---|---|
| Very easy | Easy | Neutral | Difficult | Very difficult | Not applicable |

| | **Yes** | **No** | **Not applicable** |
|---|---|---|---|
| **16.** Did you have access to a smartphone (or other device) to use the digital app? *(please tick. If you didn't use an app please choose not applicable)* | ☐ | ☐ | ☐ |
| **17.** Were you happy to record using a digital app? *(please tick. If you didn't use an app please choose not applicable)* | ☐ | ☐ | ☐ |

**18.** How did you find using the digital app? *(please circle one response. If you didn't use an app please choose not applicable)*

| | | | | | |
|---|---|---|---|---|---|
| Very easy | Easy | Neutral | Difficult | Very difficult | Not applicable |

| | Yes | No | Not applicable |
|---|---|---|---|
| **19.** Did you have anyone to support you with taking and recording readings, if needed? *(please tick)* | ☐ | ☐ | ☐ |

| | Yes | No | Not sure | Not applicable |
|---|---|---|---|---|
| **20.** Did you receive any medications as part of the COVID care at home service? *(this question may only be applicable for those who have been discharged from hospital)* | ☐ | ☐ | ☐ | ☐ |
| **21.** Were you given oxygen as part of the COVID care at home service? *(this question may only be applicable for those who have been discharged from hospital)* | ☐ | ☐ | ☐ | ☐ |

**22.** On average, how often did you (or a family member/friend) speak to a member of the COVID care at home team while receiving the service? *(please circle one response)*

| | | | | | |
|---|---|---|---|---|---|
| Several times a day | Once a day | Several times a week | Once a week | Less than once a week | Not at all |

**23.** How would you rate your contact with the COVID care at home team? *(please circle one response)*

| | | | | | |
|---|---|---|---|---|---|
| Excellent | Good | Neutral | Poor | Very poor | Not applicable |

**24.** What activities were you asked to do as part of your COVID care at home? *(Please select all that apply)*

- ☐ Take readings using an oximeter
- ☐ Fill in a diary
- ☐ Provide readings over the phone
- ☐ Record readings in a digital app
- ☐ Check over your readings for any issues
- ☐ Seek further help due to a reading being lower than recommended threshold
- ☐ Other (please specify: ____________

| | | | | | | |
|---|---|---|---|---|---|---|
| **25.** Please rate your experience of receiving COVID care at home *(please circle one option)* | Excellent | Good | Neutral | Poor | Very poor | Not applicable |
| **26.** How reassured were you by receiving COVID care at home? *(please circle one option)* | Very reassured | Reassured | Neutral | Not very reassured | Not at all reassured | Not applicable |
| **27.** Thinking about your COVID symptoms, how helpful did you find receiving COVID care at home? *(please circle one option)* | Very helpful | Helpful | Neutral | Not very helpful | Not at all helpful | Not applicable |

**28.** Would you recommend COVID care at home to your friends and family (if in a similar situation) *(please tick)*

☐ **Yes** ☐ **No** ☐ **Not sure**

**How have you found doing the following in practice?** *(please circle one option for each question)*

| | | | | | | |
|---|---|---|---|---|---|---|
| **29.** Using the oximeter | Very easy | Easy | Neutral | Difficult | Very difficult | Not applicable |
| **30.** Recording your readings using the app or diary | Very easy | Easy | Neutral | Difficult | Very difficult | Not applicable |
| **31.** Providing readings to the service | Very easy | Easy | Neutral | Difficult | Very difficult | Not applicable |
| **32.** Seeking further help if have concerns about health (escalating care if necessary) | Very easy | Easy | Neutral | Difficult | Very difficult | Not applicable |
| **33.** Contacting a healthcare professional (if needed) | Very easy | Easy | Neutral | Difficult | Very difficult | Not applicable |
| **34.** Returning the oximeter following discharge | Very easy | Easy | Neutral | Difficult | Very difficult | Not applicable |

| | **Yes** | **No** | **Not sure** |
|---|---|---|---|
| **35.** Did you understand what would happen after being discharged from COVID care at home? *(please tick)* | ☐ | ☐ | ☐ |
| **36.** Were you asked to return the oximeter once discharged from COVID care at home? *(please tick)* | ☐ | ☐ | ☐ |

**37.** Did you experience any problems with any of the following? *(Please select all that apply)*

- ☐ Using the oximeter
- ☐ Recording your readings in the app or diary
- ☐ Providing readings to the service
- ☐ Seeking further help if you had concerns about your health
- ☐ Contacting healthcare professionals when needed
- ☐ Returning the oximeter following discharge
- ☐ Other (please specify): ____________________
- ☐ I did not experience any problems

| | **Yes** | **No** | **Not sure** | **Not applicable** |
|---|---|---|---|---|
| **38.** Did you discuss these problems with a member of the COVID care at home team? *(please tick)* | ☐ | ☐ | ☐ | ☐ |

**Yes** **No** **Not sure** **Not applicable**

**39.** Was this problem (or were these problems) resolved? *(please tick)* ☐ ☐ ☐ ☐

**40.** What helped or encouraged you to use COVID care at home? *(Please select all that apply)*

- ☐ Knowing why the service is important (e.g. for patients and/or NHS)
- ☐ Knowing what to do (e.g. how to use the oximeter, how to record readings, etc)
- ☐ Own health (e.g. other health conditions)
- ☐ Having sufficient time
- ☐ Support from family or friends to use the equipment
- ☐ Support from healthcare professionals
- ☐ Wanting to do it
- ☐ Remembering to do it
- ☐ Positive views of the service (e.g. liking it/trust)
- ☐ Other (please specify) ______
- ☐ None of the above

**41.** Was there anything that made it difficult for you to use COVID care at home? *(Please select all that apply)*

- ☐ Not knowing why the service is important
- ☐ Not knowing what to do (e.g. how to use the oximeter, how to record readings, etc)
- ☐ Own health (e.g. other health conditions)
- ☐ Not having time
- ☐ Not having the right equipment
- ☐ Lack of support from family or friends
- ☐ Lack of support from healthcare professionals
- ☐ Forgetting to do it
- ☐ Not wanting to do it
- ☐ Negative views towards the service (e.g. disliking it)
- ☐ Concerns about being monitored or data use
- ☐ Other (please specify): ______
- ☐ None of the above

**42.** What could have been changed to make it easier for you to engage with COVID care at home? *(Please describe in the box below, or leave blank if not applicable)*

**43.** Which of the following scenarios best describes your experience while receiving COVID care at home? *(Please tick all that apply)*

- ☐ Stayed at home the whole time
- ☐ Asked to go to the emergency department
- ☐ Admitted to hospital

**44.** Is there anything else you'd like to tell us about your experience of receiving COVID care at home? *(Please describe in the box below, or leave blank if not applicable)*

#### Section D. About you

**Finally, we would like to ask you some questions about yourself.**

**45.** What is your gender? *(please tick one option)*

☐ Male
☐ Female
☐ Other (please specify) ______
☐ Prefer not to say

**46.** How old are you? *(please circle one option)*

| | | | | | | | | | | | | | | |
|---|---|---|---|---|---|---|---|---|---|---|---|---|---|---|
| 18-24 years | 25-29 years | 30-34 years | 35-39 years | 40-44 years | 45-49 years | 50-54 years | 55-59 years | 60-64 years | 65-69 years | 70-74 years | 75-79 years | 80-84 years | 85+ years | Prefer not to say |

**47.** How many people do you live/co-habit with? *(please circle one option)*

| | | | | | | | | | | | |
|---|---|---|---|---|---|---|---|---|---|---|---|
| 0 | 1 | 2 | 3 | 4 | 5 | 6 | 7 | 8 | 9 | 10 or more | Prefer not to say |

**48.** Which of these best describes your living arrangement?

☐ I own my home outright
☐ I own my home with a mortgage
☐ I rent from local authority/housing association
☐ I rent privately
☐ Other (e.g. living with friends or family), please specify ____________________
☐ Prefer not to say

**49.** What is your ethnic group? (*Choose one option that best describes your ethnic group or background)*

| | | |
|---|---|---|
| **White**<br>☐ English/Welsh/Scottish/ Northern Irish/British<br>☐ Irish<br>☐ Gypsy or Irish Traveller<br>☐ Any other white background, please describe: | **Asian/Asian British**<br>☐ Indian<br>☐ Pakistani<br>☐ Bangladeshi<br>☐ Chinese<br>☐ Any other Asian background, please describe: | **Other ethnic group**<br>☐ Arab<br>☐ Any other ethnic group, please describe: |
| **Mixed/Multiple ethnic groups**<br>☐ White and Black Caribbean<br>☐ White and Black African<br>☐ White and Asian<br>☐ Any other Mixed/Multiple ethnic background, please describe: | **Black/African/Caribbean/Black British**<br>☐ African<br>☐ Caribbean<br>☐ Any other Black/African/Caribbean background, please describe: | ☐ **Prefer not to say** |

**50.** At what age did you complete your continuous full time education? *(e.g. school, college, further education) Please write your age in the box below:*

___________ years

☐ Prefer not to say

**51.** Which of these best describes your highest educational qualification? *(Please tick one option)*

☐ No formal qualification
☐ GCSE/CSE/O level or equivalent
☐ A level/AS level or equivalent
☐ Degree level or higher
☐ Other, please specify ____________________
☐ Prefer not to say

**52.** Which of these best describes your current work situation? *(Please select all that apply)*

- ☐ Working full time
- ☐ Working part time
- ☐ Self-employed
- ☐ Student in higher education
- ☐ Unemployed
- ☐ Homemaker
- ☐ Retired
- ☐ Furloughed under covid-19
- ☐ Full time carer (of dependent child or adult)
- ☐ Not in work due to poor health or disability
- ☐ Other, please specify ___________________
- ☐ Prefer not to say

**53.** This question is about your sexual orientation. Do you identify as:

- ☐ Straight/heterosexual
- ☐ Gay or lesbian
- ☐ Bisexual
- ☐ Other (please specify)
- ☐ Prefer not to say

**54.** Is English your first language?

- ☐ Yes
- ☐ No, please specify ___________________
- ☐ Prefer not to say

**55.** Prior to your current illness, were your day-to-day activities limited because of a health problem or disability which has lasted, or is expected to last, at least 12 months?

- ☐ Yes limited a lot
- ☐ Yes limited a little
- ☐ No not limited at all
- ☐ Prefer not to say
- ☐ Not sure
- ☐ Not applicable

**56.** What is your postcode? (*Please write in the box below)*

☐ Prefer not to say

**Summary**

Thank you so much for completing our survey about your experiences of receiving COVID care at home. Your responses will help us to evaluate the COVID care at home service.

#### Appendix 3. Interview topic guide

##### PATIENTS

The interview should last between 30 and 60 mins, but may take longer than this, depending on how much you would like to say. We will ask questions about how you found the experience of monitoring and recording your COVID symptoms at home, and any further advice you received – we call this "COVID care at home". We will feedback the results of this evaluation to local and national NHS and public health services, and results will be made available to the general public too.

If you do not want to answer a question, you do not have to, and if you feel uncomfortable or tired we can stop the interview at any point. Let us know if you'd like a break or to come back later. We can also carry out the interview in two halves if that is easier for you to manage. Have you got any questions before we start?

**Questions for those who have received CO@h**

| Main question | Follow up questions (prompts) |
|---|---|
| 1. Please tell me a bit about yourself, | • Do you live by yourself or with others?<br>• How long have you lived in your neighbourhood?<br>• Do you have any family or friends living close by?<br>• Is English your first language? If N, ask what is their first language. |
| **FINDING OUT ABOUT THE SERVICE**<br>2. Can you tell me the story of how you ended up being referred to COVID care at home? | a. How were you referred to the service? *(may include some referrals following positive test)*<br>b. Who did you speak to and when? (virtual/face to face? How were you involved in the assessment process?) *(for assessment/triage)*<br>c. When you were first told about the service, how was it described to you?<br>d. What were your first impressions of COVID care at home? *(prompt about whether they found it reassuring or not)*<br>a. Positive impressions?<br>b. Did you have any concerns/worries? |
| **DESCRIBING COVID CARE AT HOME**<br>3. COVID care at home is carried out slightly differently in different parts of the country. Please can you tell me about what receiving COVID care at home has involved for you? | a. What equipment were you given?<br>b. How/when was the pulse oximeter delivered to you? Can you tell me how it works?<br>c. What symptoms did you have to monitor? For each, ask how often.<br>d. How did you have to record your symptoms? (e.g. paper, digital app, telephone line)<br>e. Were you offered a choice in how you recorded your symptoms?<br>f. Who did you speak to? (& how often – monitoring)<br>g. Have family members/carers been involved? If so, how? (if relevant)<br>h. Overall, how did you feel about recording and monitoring your symptoms? *(prompt about whether they found it reassuring or not)* |

| | |
|---|---|
| | i. Did you receive any medications as part of COVID care at home (for those early discharged from hospital)?<br>a. If yes. How did you find it?<br>j. Were you given oxygen as part of the COVID care at home service?<br>a. If yes, how did you find it? |
| **INFORMATION RECEIVED ON COVID CARE AT HOME when being introduced to the service**<br><br>4 a. Did you receive information on COVID care at home **in person** from a member of the care team?<br><br>If no, move to 4b.<br><br>4b. Did you receive information on COVID care at home **over the telephone or a video call** from a member of the care team?<br><br>If no, move to 4c.<br><br>4c. Were you directed to any information **to read or watch on a website** on COVID care at home?<br><br>If no, move to 4d.<br><br>4d. Were you directed to any information **to read or watch on an app** on COVID care at home?<br><br>If no, move to 5. | a. i) If yes, what information did you receive? (*Monitoring symptoms? Using oximeter? Recording symptoms? Seeking further advice?)*<br>ii) Who gave you the information?<br>Was it in your first language?<br>iii) How easy was it to understand the information?<br>iv) Did the person you spoke to describe the readings and what they mean in relation to your everyday symptoms and experience?<br><br>b. i) If yes, what information did you receive? (*Monitoring symptoms? Using oximeter? Recording symptoms? Seeking further advice?)*<br><br>ii) Who gave you the information?<br>iii) Was it in your first language?<br>iv) How easy was it to understand the information?<br><br>c. i) If yes, were you able to access the information you needed on the website?<br>ii) If no, why not?<br>If yes, what information did you receive?<br>(*Monitoring symptoms? Using oximeter? Recording symptoms? Seeking further advice*?)<br><br>iii) Was it in your first language?<br>iv) How easy was it to understand the information?<br><br>d. i) If yes, were you able to access the information you needed on the app?<br>If no, why not?<br>If i) is yes, what information did you receive?<br>*(Monitoring symptoms? Using oximeter? Recording symptoms? Seeking further advice?)*<br><br>iii) Was it in your first language? |

| | |
|---|---|
| | iv) How easy was it to understand the information? |
| **CARRYING OUT THE MONITORING**<br><br>**OXIMETER**<br>5. How did you find using the oximeter to monitor your oxygen levels? *(may like to prompt about how often they used it)* | a. What helped you to use it?<br>b. What got in the way?<br>c. What did you do when you had problems? (prompt about type of support and usefulness)<br>What could be changed to make it easier to use the oximeter? |
| **OTHER SYMPTOMS**<br>6. Overall, how did you find monitoring your other symptoms (pulse heart rate/temperature/symptoms) at home? *(may like to prompt about how often they monitored other symptoms)* | a. What worked well?<br>b. What got in the way?<br>c. What did you do when you had problems? (prompt about type of support and usefulness)<br>d. What could be changed to make it easier to monitor your other symptoms?<br>e. Was there any parts of monitoring that you were uncertain about?<br>f. Did you seek further advice from anyone about monitoring your symptoms? Prompt – who, when, how? |
| **CARRYING OUT RECORDING**<br>7. Overall, how did you find recording your readings (blood oxygen levels/symptoms/pulse heart rate/temperature)? *(using an app / diary / both) (prompt about how often they recorded their readings)* | a. What worked well?<br>b. What got in the way?<br>c. What did you do when you had problems? (prompt about type of support and usefulness)<br>d. What could be changed to make it easier to monitor your other symptoms?<br>e. Was there any part of recording your symptoms that you were uncertain about? *(prompt about whether they felt confident)*<br>r. Did you seek further advice from anyone about recording? Prompt – who, when, how? |
| **COMMUNICATING READINGS TO MEMBER OF THE TEAM**<br>**8. How have you found sending or communicating your symptoms and readings to the COVID care at home team? (if needed)** | a. What worked well?<br>b. What got in the way?<br>c. What did you do when you had problems? (prompt about type of support and usefulness)<br>d. What could be changed to make it easier to communicate your readings to the COVID care at home team?<br>e. Was there any part of communicating your readings to a member of the team that you were uncertain about?<br>r. Did you seek further advice from anyone about communicating your readings? Prompt – who, when, how? |
| **SEEKING FURTHER ADVICE**<br>8. Have you had to seek further support and help (escalate your care) because of the readings given by your oximeter or because of other things such as a change in symptoms? | a. How did this go?<br>b. What did this involve?<br><br>c. What was your experience of being sent for further support and help (e.g. escalated or admitted to hospital)? |

| | |
|---|---|
| | d. What were you instructed to do? (i.e. dial 111, dial 999, go to A&E)?<br>e. Did you self-escalate your care (seek further health care) if necessary? Why or why not? *(prompt on how they made the decision that they needed to seek further help)*<br>f. What helped you to seek further support?<br>g. What got in the way of seeking further support?<br>h. What could be changed to make it easier? |
| **DISCHARGE**<br><br>9. What was your understanding about what would happen once you are discharged from COVID care at home? | a. How did you feel after being discharged from COVID care at home? (prompt about whether they found it reassuring or not?)<br>b. Since being discharged from COVID care at home, what other services/support have you accessed? *(If discharged from COVID care at home).*<br>i. What were these?<br>ii. How often have you had to access these? |
| **RECOMMENDATIONS**<br>10. If a friend who was in a similar position to you at the start of your illness was offered COVID care at home, would you recommend it to them<br>a) over hospital care?<br>b) over no monitoring, with the option to access usual services as needed.<br><br>11. Do you have any recommendations to improve the service? | A) If yes, why? If no, why not?<br><br>B) If yes, why? If no, why not?<br><br>If yes, what? |
| 12. Is there anything else that you would like to say about what we have talked about? | |

**Interview questions on demographic characteristics (asked at the end of the interview)**

- Patient or carer? (relationship with patient, if carer)
- Gender
- Age
- How many people do you live/cohabit with?
- Which of these best describes your living arrangement? *Please select one answer (I own my home outright, I own my home with a mortgage, I rent from local authority/housing association, I rent privately, Other (e.g. living with family/friends), prefer not to say)*
- Ethnicity of patient (and family member if applicable)
- At what age did you complete your continuous full time education? *(__ years/never went to school, do not wish to answer)*
- Which of these best describes your highest *educational* qualification? *(Please select one answer) (No formal qualification, GCSE/CSE/O level or equivalent, A level/AS level or equivalent, Degree level or higher, Other (please specify), do not wish to answer)*
- Which of these best describes your current work situation? *(please tick all that apply) (Working full time, working part time, self-employed, student in higher education, unemployed, homemaker, retired, furloughed under COVID-19, Full time carer (of dependent child or adult), not in work due to poor health or disability,, Other (if other please describe), do not wish to answer).*
- Is English your first language? *(yes/no/do not wish to answer)*
- Prior to your current illness, are your day-to-day activities limited because of a health problem or disability which has lasted, or is expected to last, at least 12 months?? (Includes problems which are due to old age.) (yes, limited a lot/yes, limited a little, no/do not wish to answer)
- Sexuality
- What is your Postcode? (optional)

#### STAFF

##### Interview topic guide – staff leads

The interview should last around 45 mins, depending on how much you would like to say. We will ask questions about how your trust/CCG has implemented COVID care at home – by this we mean patient self-monitoring and recording of COVID symptoms either pre-hospital or after early discharge from hospital (also known in some places as COVID virtual wards). We will feedback the results of this evaluation to local and national NHS and public health services, and results will be made available to the general public too.

If you do not want to answer a question, you do not have to, and if you feel uncomfortable or tired we can stop the interview at any point. Have you got any questions before we start?

| ***Main question*** | ***Follow-up questions*** |
|---|---|
| **ROLE**<br>1. Can you tell me about your current role? | a. Length of time in your current role (both in the CO@H service and their usual role)<br>b. Key responsibilities – in relation to CO@H and in the 'usual' role |
| **ORIGIN OF THE MODEL**<br><br>2. Can you tell me the story of how COVID care at home started? | <br><br>a. When did it start?<br>b. Who led its development?<br>c. Has the model been or applied to other conditions apart from Covid-19? If yes, which conditions?<br>d. When did it become fully operational?<br>e. How long did it take you to get the model up and running?<br>f. Which NHS colleagues/organisations in your area (primary care, secondary care, others) were involved with the setup and delivery of this model (NHS and non-NHS colleagues/organisations at regional and/or national level? |
| **AIMS AND GOALS OF THE MODEL**<br>2. What are the aims of the model and its main features? | a. Population served<br>b. Admission criteria<br>c. Characteristics of patient groups<br>d. Pre-hospital (referral from community), step down from ED or early discharge from hospital or some combination of these?<br>e. Where located – primary, secondary, community services, integrated care Trust<br>f. Is a digital platform being used for patients to enter their observations? Is the service being delivered 24 hours per day, 7 days per week?<br>g. Availability of other services at community or primary care level for Covid-19 relevant to testing, diagnosis, mental health |
| 3. What are the main goals/outcomes of the service/model?<br><br>*(Differentiate for each patient pathway, if they have more than one, at each site)* | a. Minimise patient mortality and morbidity<br>b. Early identification of cases of deterioration<br>c. Minimise attendance/reattendance to ED<br>d. Reduced length of stay<br>e. Other |
| **RESOURCES AND PROCESSES OF THE MODEL**<br>4. Can you talk me through the staff journey from a patient contacting their GP/ED/being discharged from hospital to being discharged from COVID care at home? | a. Application of admission criteria, and referral processes (variation by age, ethnicity, deprivation)<br>b. Patient triage<br>c. Distributing pulse oximeters to patient<br>d. Patient information and training |

| | |
|---|---|
| *(Differentiate for each patient pathway, if they have more than one, at each site)*<br><br>Only relevant for staff delivering COVID care on virtual wards:<br>Are patients provided oxygen and/or medications (e.g. dexamethasone and Low Molecular Weight Heparin (LMWH)) given to patients?<br><br>5. How were pulse oximeters purchased?<br><br>6. What is the current staffing arrangement used to deliver your model? | e. Patient monitoring (who is involved, what was monitored, and how)<br>f. Mechanisms used for patient data reporting (i.e. app, paper-based)<br>g. Tools for flagging deterioration<br>h. Escalation processes (including any criteria and thresholds, safety netting)<br>i. Patient discharge from ward<br>j. Signposting to wider services<br><br>a. What was the reasoning behind including oxygen and/or medication to patients (i.e. complexity of the patient's health condition, severity of symptoms)<br><br>a. Which pulse oximeters used (NHSEI or purchased their own)?<br>b. What proportion of pulse oximeters were returned and re-used?<br><br>*a. Number of staff/ pay band/grades*<br>*b. Rota*<br>*c. Responsibilities*<br>*d. Any new additional staff been recruited e.g. working with volunteers to delivery oximeters*<br>*e. Redeployment of staff working elsewhere within the organisation*<br>*f. Training non-clinical staff to complete patient monitoring activity*<br>*g. Impact on staff morale/job satisfaction*<br>*h. Key changes which staff have noticed in their everyday working practices/current workload* |
| **FACILITATORS AND BARRIERS OF IMPLEMENTATION**.<br><br>5. Can you describe the experience of collaborating with other colleagues/orgs to set up and deliver the model?<br><br>6.. Did you draw on learning from any existing models of home monitoring?<br><br>7. How has the planning of a Covid-19 vaccination programme impacted upon the setting up/running of the CO@H model (if any)?<br><br>8. What factors have facilitated the implementation of the model during wave 2 of the pandemic?<br><br>9. What factors have been a barrier to the implementation of the model during wave 2 of the pandemic? | a. From within your organisation?<br>b. From other parts of the country? |

| | |
|---|---|
| **ADAPTATIONS**<br>10. [If model was developed between Mar and Aug 2020] Did you adapt the model after wave 1 of the pandemic? | If yes, how and why?<br><br>*a. Patient triage/risk stratification*<br>*b. Patient information and training*<br>*c. Patient monitoring*<br>*d. Mechanisms used for patient data reporting (i.e. app, paper-based)*<br>*e. Tools for flagging deterioration*<br>*f. Escalation processes*<br>*g. Patient discharge from ward*<br>*h. Staffing model*<br><br>If no, why not? |
| **MONITORING AND EVALUATION**<br>12. What data are you collecting from patients at present to monitor service delivery?<br><br>13. Can you share your thoughts about the quality of the data being recorded?<br><br>14. Can you describe the nature of data you are collecting on patient safety concerns and Covid-19 related 'near misses'?<br><br>15. Have there been changes to the data you've collected since wave 1 e.g. protected characteristics from your patients at the point of onboarding?<br><br>16. What data or information, if any, would you have liked to have collected, but couldn't? | *a. Have these data been linked to other data sources?*<br>*b. Who is able to access these data?*<br>*c. How have these data helped you to monitor progress against your expected outcomes? How else have they used these data?*<br><br>Any concerns about missing data?<br><br>If yes, why were changes made?<br>*a. If no changes have been made, may you explain why?* |
| **IMPACT**<br>17. What impact, if any, has the introduction of the service/model had on delivery of other services within your own organisation?<br><br>18. What impact, if any, has the service/model had on tackling health inequalities and/or reaching high risk populations?<br><br>19. What impact, if any, has the service/model had on the wider health and care system? | a. Have there been any concerns about patient safety and/or near misses that have occurred since the service began? (If yes, prompt for an example)<br>b. Have there been any occasions of patients refusing treatment and/or dropping out? (If yes, prompt for an example and how it has been addressed) |
| **RECOMMENDATIONS AND SUSTAINABILITY** | *a) Sustainability of the models*<br>*b) Areas that need to be improved* |

| | |
|---|---|
| 20. What advice would you give colleagues, similar to yourself, attempting to implement similar models in other areas of the country?<br><br>21. What relationships have developed or been strengthened during the course of developing and implementing the service?<br><br>22. What resources (for examples, materials, staff training) have developed as a result of developing and implementing the service?<br><br>23. Are there any new institutional structures or policies that exist as a result of developing and implementing the service?<br><br>24. How transferable is the model to other conditions? | c) *Transferability of these models to other conditions?* |
| 25. Is there anything else important that you think we should know that I have not asked you? | |

#### Appendix 4: Data sources for each research question

| Research question | Data used to address research question | Data source used (sample) |
|---|---|---|
| RQ1: Were COVID-19 remote home monitoring services adapted at a local level to increase service reach and patient engagement? | **Patient groups facing barriers**<br>Staff identified patient groups facing barriers to accessing the services and number of sites reporting adaptation of services for service user needs<br><br>**Reach/Inclusivity**<br>Service design and delivery at a local level to increase the reach and coverage (staff perspectives)<br><br>**Engagement**<br>Local service adaptations to facilitate patient engagement and uptake (staff perspectives) | • Staff survey (service lead and delivery staff)<br>• Service lead interviews |
| RQ2: Were there disparities in patients' reports of their ability to engage with the service, and were these moderated by the modality of the service? What were the potential impacts of service adaptations on health disparities according to patients? | **Accessibility of information**<br>Patient reported understanding and helpfulness of information provided by the service.<br><br>**Achievability of tasks**<br>Patient reported ease of completing tasks (i.e. using the oximeter, recording readings, providing readings, seeking further help, contacting a healthcare professional) and any problems experienced completing the tasks<br><br>**Patient reported mode of monitoring**<br>Whether submitted readings via digital app, text, email or telephone | • Patient survey (including open text responses)<br>• Patient interviews |
| RQ3: Were there disparities in patients' reports of their experience of the service? | **Experience of the service**<br>Patient rating of the service, reassurance provided by the service and how helpful the service was perceived to be. | Patient survey |

#### Appendix 5. Demographic characteristics for patient interview respondents

| Patient characteristics | Interview participants (n=62) | |
|---|---|---|
| Gender | Female | 31 (50%) |
| | Male | 31 (50%) |
| Age | <50 | 8 (13%) |
| | 50 – 59 | 19 (31%) |
| | 60 – 69 | 26 (42%) |
| | 70 – 79 | 7 (11%) |
| | =>80 | 2 (3%) |
| Living situation | Living alone | 5 (8%) |
| | Living in household of 2 | 36 (58%) |
| | Living in household of 3 or more | 21 (34%) |
| Ethnicity | White | 50 (81%) |
| | Mixed or multiple ethnic groups | 0 (0%) |
| | Asian or Asian British | 7 (11%) |
| | Black, African, Caribbean or Black British | 3 (5%) |
| | Other ethnic group | 0 (0%) |
| | Not known | 2 (3%) |
| English first language | Yes | 54 (87%) |
| | No | 7 (11%) |
| | Not known | 1 (2%) |
| Highest level of education | No formal qualifications | 15 (24%) |
| | GCSE/CSE/O level | 22 (35%) |
| | A level | 5 (8%) |
| | Degree level or higher | 15 (24%) |
| | Not known | 5 (8%) |
| Work situation | Retired | 23 (37%) |
| | Working full-time | 25 (40%) |
| | Working part-time | 2 (3%) |
| | Furloughed | 1 (2%) |
| | Unemployed or homemaker | 3 (5%) |
| | Not working due to poor health/disability | 7 (11%) |
| | Not known | 1 (2%) |
| Health problem or disability which limits daily activities | No | 46 (74%) |
| | Limited a little | 6 (10%) |
| | Limited a lot | 6 (10%) |
| | Limited – not specified | 4 (6%) |
| Service received | CO@h | 44 (71%) |
| | Virtual ward | 13 (21%) |
| | Both | 2 (3%) |
| | Not known | 2 (3%) |
| | Not applicable – service offered but not received | 1 (2%) |
| Mode of monitoring | Analogue only | 19 (31%) |
| | Tech-enabled | 41 (66%) |
| | Not known (carers submitted readings) | 1 (2%) |
| | Not applicable – service offered but not received | 1 (2%) |

#### Appendix 6. Demographic characteristics for staff survey respondents

| | Service manager or clinical lead (n=70) | Delivery staff (n=222) | Total (n=292) |
|---|---|---|---|
| **Professional role within the service* n (%)** | | | |
| Clinical | 49 (70) | 157 (71) | 206 (71) |
| Non-clinical | 23 (33) | 63 (28) | 86 (29) |
| Volunteer | 0 | 0 | 0 |
| Student | 1 (1) | 4 (2) | 5 (2) |
| Other | 4 (6) | | 4 (1) |
| **Redeployed n (%)** | | | |
| Yes | 7 (10) | 71 (32) | 78 (27) |
| No | 52 (74) | 131 (59) | 183 (63) |
| Not applicable | 11 (16) | 20 (9) | 31 (11) |
| **Numbers redeployed due to shielding** n (%)** | | | |
| Yes | 0 | 21 (30) | 21 (27) |
| No | 7 (100) | 50 (70) | 57 (73) |
| **Sharing Covid role with any other roles n (%)** | | | |
| Yes | 52 (74) | 137 (62) | 189 (65) |
| No | 13 (19) | 78 (35) | 91 (31) |
| Not applicable | 5 (7) | 7 (3) | 12 (4) |
| **Length of time involved in the service n (%)** | | | |
| < 1 month | 1 (1) | 13 (6) | 14 (5) |
| 1-3 months | 14 (20) | 91 (41) | 105 (36) |
| 4-6 months | 37 (53) | 81 (37) | 118 (40) |
| 7-9 months | 13 (19) | 18 (8) | 31 (11) |
| > 10 months | 5 (7) | 19 (9) | 24 (8) |
| **Where based for the service n (%)** | | | |
| Remotely | 3 (43) | 106 (48) | 136 (47) |
| Hot hub | 8 (11) | 23 (10) | 31 (11) |
| GP practice | 6 (9) | 7 (3) | 13 (4) |
| Hospital | 15 (21) | 32 (14) | 47 (16) |
| Other | 11 (16) | 54 (24) | 65 (22) |

Note. Service leads n=70 and delivering staff n=222 unless specified
*Respondents were able to select more than one response option
**Service lead n=7, delivery staff n=71

#### Appendix 7. Patient survey analysis

**Table 1.** Fundamental factors and patient engagement with and experience of the service

| | **Gender** | | **Age** | | | | **Ethnicity** | | **Health status** | |
|---|---|---|---|---|---|---|---|---|---|---|
| | **Male N (%)** | **Female N (%)** | **Under 50 years *N (%)*** | **50-64 years *N (%)*** | **65-79 years *N (%)*** | **80 years and over *N (%)*** | **White ethnic group *N (%)*** | **Minority ethnic group *N (%)*** | **Limited a little or a lot *N (%)*** | **Not limited at all *N (%)*** |
| **Understanding information** | | | | | | | | | | |
| Very easy | 234 (61.9) | 333 (64.3) | 121 (63.4) | 286 (67.1) | 149 (60.3) | 15 (39.5) | 527 (64.9) | 35 (48.6) | 196 (58.5) | 318 (66.8) |
| Easy | 123 (32.5) | 152 (29.3) | 62 (32.5) | 116 (27.2) | 80 (32.4) | 19 (50.0) | 246 (30.3) | 23 (31.9) | 115 (34.3) | 133 (27.9) |
| Neutral | 18 (4.8) | 27 (5.2) | 8 (4.2) | 19 (4.5) | 14 (5.7) | 4 (10.5) | 34 (4.2) | 11 (15.3) | 19 (5.7) | 22 (4.6) |
| Difficult | 3 (0.8) | 5 (1.0) | 0 | 4 (0.9) | 4 (1.6) | 0 | 5 (0.6) | 3 (4.2) | 5 (1.5) | 3 (0.6) |
| Very difficult | 0 | 1 (0.2) | 0 | 1 (0.2) | 0 | 0 | 0 | 0 | 0 | 0 |
| *Total* | 378 | 518 | 191 | 426 | 247 | 38 | 812 | 72 | 335 | 476 |
| **P-value** | p=0.561[1] | | **p=0.005**[2] | | | | **p=0.001**[1] | | p=0.014[1] | |
| **How helpful was the information** | | | | | | | | | | |
| Very helpful | 238 (64.0) | 359 (70.0) | 129 (68.3) | 296 (71.0) | 163 (66.0) | 16 (42.1) | 548 (68.5) | 42 (57.5) | 218 (65.7) | 322 (68.8) |
| Helpful | 113 (30.4) | 129 (25.1) | 53 (28.0) | 99 (23.7) | 70 (28.3) | 19 (50.0) | 216 (27.0) | 22 (30.1) | 94 (28.3) | 123 (26.3) |
| Neutral | 18 (4.8) | 18 (3.5) | 6 (3.2) | 18 (4.3) | 9 (3.6) | 3 (7.9) | 30 (3.8) | 6 (8.2) | 18 (5.4) | 17 (3.6) |
| Not very helpful | 2 (0.5) | 4 (0.8) | 1 (0.5) | 2 (0.5) | 3 (1.2) | 0 | 5 (0.6) | 1 (1.4) | 1 (0.3) | 4 (0.9) |
| Not at all helpful | 1 (0.3) | 3 (0.6) | 0 | 2 (0.5) | 2 (0.8) | 0 | 1 (0.1) | 2 (2.7) | 1 (0.3) | 2 (0.4) |
| *Total* | 372 | 513 | 189 | 417 | 247 | 38 | 800 | 73 | 332 | 468 |
| **P-value** | p=0.067[1] | | **p=0.005**[2] | | | | p=0.024[1] | | p=0.334[1] | |
| **Achievability of service tasks** | | | | | | | | | | |
| **Using the oximeter** | | | | | | | | | | |
| Very easy | 296 (77.5) | 430 (82.5) | 156 (82.1) | 348 (82.5) | 199 (78.3) | 28 (65.1) | 666 (81.1) | 50 (71.4) | 261 (77.2) | 397 (83.1) |
| Easy | 76 (19.9) | 81 (15.5) | 33 (17.4) | 64 (15.2) | 46 (18.1) | 15 (34.9) | 141 (17.2) | 15 (21.4) | 70 (20.7) | 71 (14.9) |
| Neutral | 7 (1.8) | 7 (1.3) | 1 (0.5) | 8 (1.9) | 5 (2.0) | 0 | 9 (1.1) | 5 (7.1) | 3 (0.9) | 9 (1.9) |
| Difficult | 3 (0.8) | 1 (0.2) | 0 | 1 (0.2) | 3 (1.2) | 0 | 4 (0.5) | 0 | 4 (1.2) | 0 |
| Very difficult | 0 | 2 (0.4) | 0 | 1 (0.2) | 1 (0.4) | 0 | 1 (0.1) | 0 | 0 | 1 (0.2) |
| *Total* | 382 | 521 | 190 | 422 | 254 | 43 | 821 | 70 | 338 | 478 |
| **P-value** | p=0.059[1] | | p=0.044[2] | | | | p=0.034[1] | | p=0.042[1] | |
| **Recording readings** | | | | | | | | | | |
| Very easy | 249 (71.3) | 374 (78.9) | 144 (80.0) | 291 (76.8) | 173 (73.6) | 18 (54.5) | 577 (77.3) | 39 (59.1) | 210 (70.0) | 349 (79.3) |
| Easy | 89 (25.5) | 87 (18.4) | 32 (17.8) | 77 (20.3) | 54 (23.0) | 15 (45.5) | 152 (20.4) | 21 (31.8) | 81 (27.0) | 80 (18.2) |
| Neutral | 8 (2.3) | 7 (1.5) | 3 (1.7) | 7 (1.8) | 5 (2.1) | 0 | 9 (1.2) | 6 (9.1) | 6 (2.0) | 8 (1.8) |
| Difficult | 3 (0.9) | 5 (1.1) | 1 (0.6) | 3 (0.8) | 3 (1.3) | 0 | 8 (1.1) | 0 | 3 (1.0) | 3 (0.7) |
| Very difficult | 0 | 1 (0.2) | 0 | 1 (0.3) | 0 | 0 | 0 | 0 | 0 | 0 |

| | | | | | | | | | | |
|---|---|---|---|---|---|---|---|---|---|---|
| *Total* | 349 | 474 | 180 | 379 | 235 | 33 | 746 | 66 | 300 | 440 |
| **P-value** | p=0.015[1] | | p=0.022[2] | | | | **p<0.001**[1] | | **p=0.004**[1] | |
| **Providing readings** | | | | | | | | | | |
| Very easy | 253 (70.1) | 409 (80.0) | 149 (78.8) | 321 (78.3) | 176 (72.7) | 21 (56.8) | 609 (76.9) | 44 (64.7) | 235 (71.4) | 359 (78.6) |
| Easy | 95 (26.3) | 91 (17.8) | 36 (19.0) | 80 (19.5) | 56 (23.1) | 16 (43.2) | 167 (21.1) | 18 (26.5) | 82 (24.9) | 91 (19.9) |
| Neutral | 11 (3.0) | 8 (1.6) | 3 (1.6) | 8 (2.0) | 8 (3.3) | 0 | 12 (1.5) | 6 (8.8) | 11 (3.3) | 5 (1.1) |
| Difficult | 2 (0.6) | 1 (0.2) | 1 (0.5) | 0 | 1 (0.4) | 0 | 3 (0.4) | 0 | 1 (0.3) | 1 (0.2) |
| Very difficult | 0 | 2 (0.4) | 0 | 1 (0.2) | 1 (0.4) | 0 | 1 (0.1) | 0 | 0 | 1 (0.2) |
| *Total* | 361 | 511 | 189 | 410 | 242 | 37 | 792 | 68 | 329 | 457 |
| **P-value** | **p=0.001**[1] | | p=0.016[2] | | | | p=0.013[1] | | p=0.017[1] | |
| **Seeking further help** | | | | | | | | | | |
| Very easy | 177 (59.0) | 269 (62.4) | 110 (65.9) | 219 (61.5) | 111 (60.0) | 13 (41.9) | 408 (62.5) | 35 (50.0) | 161 (57.3) | 242 (63.0) |
| Easy | 82 (27.3) | 106 (24.6) | 35 (21.0) | 91 (25.6) | 48 (25.9) | 15 (48.4) | 165 (25.3) | 18 (25.7) | 79 (28.1) | 93 (24.2) |
| Neutral | 30 (10.0) | 33 (7.7) | 16 (9.6) | 32 (9.0) | 14 (7.6) | 1 (3.2) | 52 (8.0) | 11 (15.7) | 22 (7.8) | 34 (8.9) |
| Difficult | 10 (3.3) | 17 (3.9) | 3 (1.8) | 12 (3.4) | 11 (5.9) | 1 (3.2) | 22 (3.4) | 6 (8.6) | 14 (5.0) | 14 (3.6) |
| Very difficult | 1 (0.3) | 6 (1.4) | 3 (1.8) | 2 (0.6) | 1 (0.5) | 1 (3.2) | 6 (0.9) | 0 | 5 (1.8) | 1 (0.3) |
| *Total* | 300 | 431 | 167 | 356 | 185 | 31 | 653 | 70 | 281 | 384 |
| **P-value** | p=0.437[1] | | p=0.216[2] | | | | p=0.015[1] | | p=0.128[1] | |
| **Contacting HCP** | | | | | | | | | | |
| Very easy | 152 (49.4) | 250 (58.0) | 97 (58.4) | 197 (55.2) | 99 (52.9) | 13 (39.4) | 365 (55.6) | 33 (47.1) | 143 (50.0) | 215 (56.0) |
| Easy | 99 (32.1) | 114 (26.5) | 42 (25.3) | 101 (28.3) | 57 (30.5) | 13 (39.4) | 188 (28.6) | 20 (28.6) | 85 (29.7) | 111 (28.9) |
| Neutral | 32 (10.4) | 43 (10.0) | 18 (10.8) | 38 (10.6) | 16 (8.6) | 3 (9.1) | 63 (9.6) | 10 (14.3) | 32 (11.2) | 37 (9.6) |
| Difficult | 22 (7.1) | 17 (3.9) | 7 (4.2) | 16 (4.5) | 14 (7.5) | 2 (6.1) | 32 (4.9) | 7 (10.0) | 21 (7.3) | 18 (4.7) |
| Very difficult | 3 (1.0) | 7 (1.6) | 2 (1.2) | 5 (1.4) | 1 (0.5) | 2 (6.1) | 9 (1.4) | 0 | 5 (1.7) | 3 (0.8) |
| *Total* | 308 | 431 | 166 | 357 | 187 | 33 | 657 | 70 | 286 | 384 |
| **P-value** | **p=0.025**[1] | | p=0.268[2] | | | | p=0.100[1] | | p=0.061[1] | |
| **Rating of the service** | | | | | | | | | | |
| Excellent | 247 (65.5) | 360 (69.2) | 137 (71.0) | 295 (69.7) | 159 (64.9) | 22 (52.4) | 555 (68.2) | 50 (70.4) | 228 (67.1) | 325 (68.9) |
| Good | 106 (28.1) | 126 (24.2) | 48 (24.9) | 101 (23.9) | 69 (28.2) | 13 (31.0) | 206 (25.3) | 18 (25.4) | 85 (25.0) | 121 (25.6) |
| Neutral | 21 (5.6) | 28 (5.4) | 8 (4.1) | 23 (5.4) | 13 (5.3) | 6 (14.3) | 46 (5.7) | 2 (2.8) | 25 (7.4) | 20 (4.2) |
| Poor | 2 (0.5) | 4 (0.8) | 0 | 3 (0.7) | 3 (1.2) | 0 | 5 (0.6) | 1 (1.4) | 2 (0.6) | 4 (0.8) |
| Very poor | 1 (0.3) | 2 (0.4) | 0 | 1 (0.2) | 1 (0.4) | 1 (2.4) | 2 (0.2) | 0 | 0 | 2 (0.4) |
| *Total* | 377 | 520 | 193 | 423 | 245 | 42 | 814 | 71 | 340 | 472 |
| **P-value** | p=0.289[1] | | p=0.034[2] | | | | p=0.634[1] | | p= 0.478[1] | |
| **How helpful was the service** | | | | | | | | | | |
| Very helpful | 225 (60.3) | 339 (66.3) | 129 (67.2) | 277 (66.7) | 145 (60.2) | 18 (42.9) | 515 (64.2) | 47 (65.3) | 213 (63.8) | 291 (62.3) |
| Helpful | 105 (28.2) | 128 (25.0) | 47 (24.5) | 104 (25.1) | 66 (27.4) | 16 (38.1) | 207 (25.8) | 20 (27.8) | 86 (25.7) | 131 (28.1) |
| Neutral | 33 (8.8) | 29 (5.7) | 13 (6.8) | 24 (5.8) | 20 (8.3) | 5 (11.9) | 58 (7.2) | 2 (2.8) | 26 (7.8) | 30 (6.4) |
| Not very helpful | 5 (1.3) | 9 (1.8) | 2 (1.0) | 7 (1.7) | 6 (2.5) | 0 | 13 (1.6) | 2 (2.8) | 8 (2.4) | 7 (1.5) |
| Not at all helpful | 5 (1.3) | 6 (1.2) | 1 (0.5) | 3 (0.7) | 4 (1.7) | 3 (7.1) | 9 (1.1) | 1 (1.4) | 1 (0.3) | 8 (1.7) |
| *Total* | 373 | 511 | 192 | 415 | 241 | 42 | 802 | 72 | 334 | 467 |
| **P-value** | p=0.056[1] | | **p=0.004**[2] | | | | p=0.762[1] | | p=0.515[1] | |

| | | | | | | | | | | |
|---|---|---|---|---|---|---|---|---|---|---|
| **How reassuring was the service** | | | | | | | | | | |
| Very reassured | 233 (61.8) | 350 (67.7) | 128 (66.7) | 279 (66.1) | 156 (63.7) | 25 (61.0) | 532 (65.6) | 46 (64.8) | 218 (64.7) | 312 (66.0) |
| Reassured | 112 (29.7) | 125 (24.2) | 51 (26.6) | 112 (26.5) | 66 (26.9) | 9 (22.0) | 217 (26.8) | 17 (23.9) | 84 (24.9) | 128 (27.1) |
| Neutral | 28 (7.4) | 25 (4.8) | 12 (6.3) | 20 (4.7) | 15 (6.1) | 6 (14.6) | 45 (5.5) | 4 (5.6) | 26 (7.7) | 21 (4.4) |
| Not very reassured | 3 (0.8) | 11 (2.1) | 0 | 9 (2.1) | 4 (1.6) | 1 (2.4) | 12 (1.5) | 3 (4.2) | 8 (2.4) | 7 (1.5) |
| Not at all reassured | 1 (0.3) | 6 (1.2) | 1 (0.5) | 2 (0.5) | 4 (1.6) | 0 | 5 (0.6) | 1 (1.4) | 1 (0.3) | 5 (1.1) |
| *Total* | 377 | 517 | 192 | 422 | 245 | 41 | 811 | 71 | 337 | 473 |
| **P-value** | p=0.104[1] | | p=0.640[2] | | | | p=0.715[1] | | p=0.753[1] | |

Note. Bold text indicates significance at p< .05 level.

p-values are derived from Mann-Whitney U test[1] or Kruskal-Wallis H test[2]

**Table 2.** Socioeconomic factors and patient engagement with and experience of the service

| | **Level of education** | | | **Employment status** | | **First language** | | **Living situation** | |
|---|---|---|---|---|---|---|---|---|---|
| | **No formal qualificatio n** *N (%)* | **GCSE level or equivalent** *N (%)* | **AS, A level, degree level or equivalent** *N (%)* | **Full-time, part-time or self-employed** *N (%)* | **Not in employment** *N (%)* | **English first language** *N (%)* | **Other** *N (%)* | **Living alone** *N (%)* | **Living with others** *N (%)* |
| **Understanding information** | | | | | | | | | |
| Very easy | 86 (60.1) | 178 (67.7) | 202 (65.2) | 325 (66.5) | 238 (60.6) | 532 (63.9) | 30 (51.7) | 73 (56.6) | 477 (64.3) |
| Easy | 51 (35.7) | 74 (28.1) | 89 (28.7) | 142 (29.0) | 129 (32.8) | 254 (30.5) | 22 (37.9) | 49 (38.0) | 219 (29.5) |
| Neutral | 5 (3.5) | 11 (4.2) | 15 (4.8) | 20 (4.1) | 21 (5.3) | 39 (4.7) | 6 (10.3) | 5 (3.9) | 39 (5.3) |
| Difficult | 1 (0.7) | 0 | 4 (1.3) | 2 (0.4) | 5 (1.3) | 8 (1.0) | 0 | 1 (0.8) | 7 (0.9) |
| Very difficult | 0 | 0 | 0 | 0 | 0 | 0 | 0 | 1 (0.8) | 0 |
| *Total* | 143 | 263 | 310 | 489 | 393 | 833 | 58 | 129 | 742 |
| **P-value** | p=0.352[2] | | | p=0.053[1] | | p=0.050[1] | | p=0.134[1] | |
| **How helpful was the information** | | | | | | | | | |
| Very helpful | 100 (71.4) | 187 (72.2) | 201 (65.3) | 338 (70.4) | 255 (65.2) | 561 (68.3) | 35 (59.3) | 82 (64.6) | 494 (67.3) |
| Helpful | 35 (25.0) | 66 (25.5) | 83 (26.9) | 120 (25.0) | 116 (29.7) | 219 (26.7) | 19 (32.2) | 38 (29.9) | 201 (27.4) |
| Neutral | 4 (2.9) | 5 (1.9) | 18 (5.8) | 19 (4.0) | 15 (3.8) | 32 (3.9) | 5 (8.5) | 3 (2.4) | 33 (4.5) |
| Not very helpful | 1 (0.7) | 1 (0.4) | 4 (1.3) | 3 (0.6) | 3 (0.8) | 6 (0.7) | 0 | 2 (1.6) | 4 (0.5) |
| Not at all helpful | 0 | 0 | 2 (0.6) | 0 | 2 (0.5) | 3 (0.4) | 0 | 2 (1.6) | 2 (0.3) |
| *Total* | 140 | 259 | 308 | 480 | 391 | 821 | 59 | 127 | 734 |
| **P-value** | p=0.080[2] | | | p=0.109[1] | | p=0.133[1] | | p=0.543[1] | |
| **Achievability of service tasks** | | | | | | | | | |
| **Using the oximeter** | | | | | | | | | |
| Very easy | 115 (80.4) | 216 (82.1) | 261 (82.3) | 404 (82.8) | 315 (78.8) | 675 (80.5) | 44 (75.9) | 103 (76.9) | 604 (81.0) |
| Easy | 28 (19.6) | 44 (16.7) | 45 (14.2) | 76 (15.6) | 74 (18.5) | 146 (17.4) | 13 (22.4) | 27 (20.1) | 126 (16.9) |
| Neutral | 0 | 1 (0.4) | 9 (2.8) | 8 (1.6) | 6 (1.5) | 13 (1.5) | 1 (1.7) | 2 (1.5) | 12 (1.6) |
| Difficult | 0 | 1 (0.4) | 2 (0.6) | 0 | 4 (1.0) | 4 (0.5) | 0 | 0 | 4 (0.5) |
| Very difficult | 0 | 1 (0.4) | 0 | 0 | 1 (0.3) | 1 (0.1) | 0 | 2 (1.5) | 0 |
| *Total* | 143 | 263 | 317 | 488 | 400 | 839 | 58 | 134 | 746 |
| **P-value** | p=0.931[2] | | | p=0.115[1] | | p=0.418[1] | | p=0.262[1] | |
| **Recording readings** | | | | | | | | | |
| Very easy | 95 (75.4) | 190 (79.5) | 223 (76.6) | 359 (79.1) | 257 (72.8) | 580 (76.3) | 37 (67.3) | 79 (68.1) | 526 (76.5) |
| Easy | 30 (23.8) | 44 (18.4) | 55 (18.9) | 82 (18.1) | 86 (24.4) | 159 (20.9) | 16 (29.1) | 31 (26.7) | 144 (20.9) |
| Neutral | 0 | 3 (1.3) | 9 (3.1) | 9 (2.0) | 6 (1.7) | 13 (1.7) | 2 (3.6) | 1 (0.9) | 14 (2.0) |
| Difficult | 1 (0.8) | 2 (0.8) | 4 (1.4) | 4 (0.9) | 4 (1.1) | 8 (1.1) | 0 | 4 (3.4) | 4 (0.6) |

| | | | | | | | | | |
|---|---|---|---|---|---|---|---|---|---|
| Very difficult | 0 | 0 | 0 | 0 | 0 | 0 | 0 | 1 (0.9) | 0 |
| *Total* | 126 | 239 | 291 | 454 | 353 | 760 | 55 | 116 | 688 |
| **P-value** | p=0.600[2] | | | p=0.044[1] | | p=0.136[1] | | p=0.043[1] | |
| **Providing readings** | | | | | | | | | |
| Very easy | 105 (76.1) | 196 (77.5) | 231 (75.2) | 378 (79.1) | 278 (73.2) | 618 (76.3) | 39 (69.6) | 92 (70.8) | 553 (76.7) |
| Easy | 30 (21.7) | 53 (20.9) | 64 (20.8) | 91 (19.0) | 90 (23.7) | 172 (21.2) | 14 (25.0) | 33 (25.4) | 149 (20.7) |
| Neutral | 3 (2.2) | 4 (1.6) | 9 (2.9) | 7 (1.5) | 10 (2.6) | 16 (2.0) | 3 (5.4) | 3 (2.3) | 16 (2.2) |
| Difficult | 0 | 0 | 2 (0.7) | 2 (0.4) | 1 (0.3) | 3 (0.4) | 0 | 1 (0.8) | 2 (0.3) |
| Very difficult | 0 | 0 | 1 (0.3) | 0 | 1 (0.3) | 1 (0.1) | 0 | 1 (0.8) | 1 (0.1) |
| *Total* | 138 | 253 | 307 | 478 | 380 | 810 | 56 | 130 | 721 |
| **P-value** | p=0.753[2] | | | p=0.038[1] | | p=0.229[1] | | p=0.138[1] | |
| **Seeking further help** | | | | | | | | | |
| Very easy | 76 (62.3) | 146 (68.5) | 146 (57.7) | 277 (65.3) | 165 (55.9) | 410 (61.1) | 32 (58.2) | 59 (54.1) | 374 (62.0) |
| Easy | 39 (32.0) | 48 (22.5) | 63 (24.9) | 101 (23.8) | 84 (28.5) | 172 (25.6) | 16 (29.1) | 33 (30.3) | 152 (25.2) |
| Neutral | 4 (3.3) | 16 (7.5) | 23 (9.1) | 32 (7.5) | 28 (9.5) | 56 (8.3) | 6 (10.9) | 7 (6.4) | 53 (8.8) |
| Difficult | 3 (2.5) | 2 (0.9) | 19 (7.5) | 13 (3.1) | 14 (4.7) | 28 (4.2) | 0 | 6 (5.5) | 21 (3.5) |
| Very difficult | 0 | 1 (0.5) | 2 (0.8) | 1 (0.2) | 4 (1.4) | 5 (0.7) | 1 (1.8) | 4 (3.7) | 3 (0.5) |
| *Total* | 122 | 213 | 253 | 424 | 295 | 671 | 55 | 109 | 603 |
| **P-value** | p=0.015[2] | | | **p=0.007**[1] | | p=0.775[1] | | p=0.100[1] | |
| **Contacting HCP** | | | | | | | | | |
| Very easy | 72 (61.5) | 127 (58.3) | 131 (51.4) | 244 (58.5) | 154 (50.2) | 366 (54.2) | 31 (55.4) | 59 (55.1) | 328 (53.7) |
| Easy | 36 (30.8) | 59 (27.1) | 73 (28.6) | 113 (27.1) | 98 (31.9) | 197 (29.2) | 15 (26.8) | 28 (26.2) | 181 (29.6) |
| Neutral | 5 (4.3) | 20 (9.2) | 29 (11.4) | 40 (9.6) | 30 (9.8) | 67 (9.9) | 7 (12.5) | 12 (11.2) | 62 (10.1) |
| Difficult | 4 (3.4) | 9 (4.1) | 20 (7.8) | 19 (4.6) | 18 (5.9) | 37 (5.5) | 2 (3.6) | 5 (4.7) | 33 (5.4) |
| Very difficult | 0 | 3 (1.4) | 2 (0.8) | 1 (0.2) | 7 (2.3) | 8 (1.2) | 1 (1.8) | 3 (2.8) | 7 (1.1) |
| *Total* | 117 | 218 | 255 | 417 | 307 | 675 | 56 | 107 | 611 |
| **P-value** | p=0.036[2] | | | p=0.021[1] | | p=0.933[1] | | p=0.976[1] | |
| **Rating of the service** | | | | | | | | | |
| Excellent | 109 (75.9) | 172 (66.7) | 208 (66.2) | 356 (72.7) | 247 (63.2) | 564 (67.8) | 40 (67.8) | 88 (67.2) | 500 (67.5) |
| Good | 29 (20.1) | 69 (26.7) | 84 (26.8) | 114 (23.3) | 111 (28.4) | 213 (25.6) | 16 (27.1) | 34 (26.0) | 194 (26.2) |
| Neutral | 5 (3.5) | 16 (6.2) | 16 (5.1) | 19 (3.9) | 26 (6.6) | 47 (5.6) | 3 (5.1) | 7 (5.3) | 40 (5.4) |
| Poor | 1 (0.7) | 1 (0.4) | 4 (1.3) | 1 (0.2) | 5 (1.3) | 6 (0.7) | 0 | 0 | 6 (0.8) |
| Very poor | 0 | 0 | 2 (0.6) | 0 | 2 (0.5) | 2 (0.2) | 0 | 2 (1.5) | 1 (0.1) |
| *Total* | 144 | 258 | 314 | 490 | 391 | 832 | 59 | 131 | 741 |
| **P-value** | p=0.094[2] | | | **p=0.001**[1] | | p=0.932[1] | | p=0.912[1] | |

| | | | | | | | | | |
|---|---|---|---|---|---|---|---|---|---|
| **How helpful was the service** | | | | | | | | | |
| Very helpful | 99 (69.2) | 164 (63.8) | 183 (59.6) | 326 (68.1) | 232 (59.6) | 524 (63.9) | 34 (58.6) | 83 (63.4) | 467 (64.1) |
| Helpful | 36 (25.2) | 71 (27.6) | 88 (28.7) | 120 (25.1) | 108 (27.8) | 214 (26.1) | 18 (31.0) | 37 (28.2) | 189 (25.1) |
| Neutral | 6 (4.2) | 17 (6.6) | 23 (7.5) | 28 (5.8) | 30 (7.7) | 59 (7.2) | 4 (6.9) | 7 (5.3) | 51 (7.0) |
| Not very helpful | 2 (1.4) | 5 (1.9) | 7 (2.3) | 3 (0.6) | 12 (3.1) | 13 (1.6) | 2 (3.4) | 0 | 15 (2.1) |
| Not at all helpful | 0 | 0 | 6 (2.0) | 2 (0.4) | 7 (1.8) | 10 (1.2) | 0 | 4 (3.1) | 7 (1.0) |
| *Total* | 143 | 257 | 307 | 479 | 389 | 820 | 58 | 131 | 729 |
| **P-value** | p=0.077[2] | | | **p=0.003**[1] | | p=0.464[1] | | p=0.972[1] | |
| **How reassuring was the service** | | | | | | | | | |
| Very reassured | 101 (70.6) | 170 (66.1) | 201 (64.0) | 332 (68.0) | 245 (62.8) | 542 (65.2) | 35 (61.4) | 90 (68.2) | 475 (64.5) |
| Reassured | 34 (23.8) | 72 (28.0) | 80 (25.5) | 129 (26.4) | 102 (26.2) | 219 (26.4) | 17 (29.8) | 31 (23.5) | 203 (27.5) |
| Neutral | 7 (4.9) | 10 (3.9) | 22 (7.0) | 22 (4.5) | 28 (7.2) | 50 (6.0) | 4 (7.0) | 7 (5.3) | 42 (5.7) |
| Not very reassured | 1 (0.7) | 5 (1.9) | 6 (1.9) | 4 (0.8) | 10 (2.6) | 14 (1.7) | 1 (1.8) | 0 | 14 (1.9) |
| Not at all reassured | 0 | 0 | 5 (1.6) | 1 (0.2) | 5 (1.3) | 6 (0.7) | 0 | 4 (3.0) | 3 (0.4) |
| *Total* | 143 | 257 | 314 | 488 | 390 | 831 | 57 | 132 | 737 |
| **P-value** | p=0.255[2] | | | p=0.038[1] | | p=0.594[1] | | p=0.484[1] | |

Note. Bold text indicates significance at $p < .05$ level.

p values are derived from Mann-Whitney U test[1] or Kruskal-Wallis H test[2]

**Table 3.** Geographic factors and patient engagement with and experience of the service

| | Deprivation score Deciles* | | | | |
|---|---|---|---|---|---|
| | **1&2 (most deprived)** *N (%)* | **3&4** *N (%)* | **5&6** *N (%)* | **7&8** *N (%)* | **9&10 (least deprived)** *N (%)* |
| **Understanding information** | | | | | |
| Very easy | 113 (62.4) | 80 (59.7) | 93 (63.7) | 107 (67.3) | 92 (69.2) |
| Easy | 58 (32.0) | 46 (34.3) | 43 (29.5) | 46 (28.9) | 37 (27.8) |
| Neutral | 10 (5.5) | 6 (4.5) | 7 (4.8) | 5 (3.1) | 4 (3.0) |
| Difficult | 0 | 2 (1.5) | 3 (2.1) | 1 (0.6) | 0 |
| Very difficult | 0 | 0 | 0 | 0 | 0 |
| *Total* | 181 | 134 | 146 | 159 | 133 |
| **P-value** | p=0.404² | | | | |
| **How helpful was the information** | | | | | |
| Very helpful | 122 (68.5) | 86 (64.7) | 100 (68.0) | 109 (69.9) | 95 (72.5) |
| Helpful | 47 (26.4) | 41 (30.8) | 35 (23.8) | 43 (27.6) | 31 (23.7) |
| Neutral | 8 (4.5) | 4 (3.0) | 9 (6.1) | 4 (2.6) | 3 (2.3) |
| Not very helpful | 1 (0.6) | 1 (0.8) | 2 (1.4) | 0 | 2 (1.5) |
| Not at all helpful | 0 | 1 (0.8) | 1 (0.7) | 0 | 0 |
| *Total* | 178 | 133 | 147 | 156 | 131 |
| **P-value** | p=0.695² | | | | |
| **Achievability of service tasks** | | | | | |
| **Using the oximeter** | | | | | |
| Very easy | 132 (74.2) | 108 (80.0) | 130 (87.8) | 134 (83.8) | 116 (84.1) |
| Easy | 43 (24.2) | 23 (17.0) | 14 (9.5) | 23 (14.4) | 20 (14.5) |
| Neutral | 3 (1.7) | 3 (2.2) | 3 (2.0) | 2 (1.3) | 2 (1.4) |
| Difficult | 0 | 0 | 1 (0.7) | 1 (0.6) | 0 |
| Very difficult | 0 | 1 (0.7) | 0 | 0 | 0 |
| *Total* | 178 | 135 | 148 | 160 | 138 |
| **P-value** | p=0.026² | | | | |
| **Recording readings** | | | | | |
| Very easy | 110 (70.1) | 94 (76.4) | 110 (80.3) | 118 (81.4) | 98 (79.7) |
| Easy | 40 (25.5) | 23 (18.7) | 26 (19.0) | 24 (16.6) | 21 (17.1) |
| Neutral | 4 (2.5) | 4 (3.3) | 1 (0.7) | 3 (2.1) | 2 (1.6) |
| Difficult | 3 (1.9) | 2 (1.6) | 0 | 0 | 2 (1.6) |
| Very difficult | 0 | 0 | 0 | 0 | 0 |
| *Total* | 157 | 123 | 137 | 145 | 123 |

| | | | | | |
|---|---|---|---|---|---|
| **P-value** | p=0.107[2] | | | | |
| **Providing readings** | | | | | |
| Very easy | 127 (73.8) | 103 (76.9) | 111 (77.6) | 122 (79.2) | 103 (78.6) |
| Easy | 39 (22.7) | 25 (18.7) | 29 (20.3) | 29 (18.8) | 26 (19.8) |
| Neutral | 5 (2.9) | 5 (3.7) | 3 (2.1) | 3 (1.9) | 0 |
| Difficult | 1 (0.6) | 1 (0.7) | 0 | 0 | 1 (0.8) |
| Very difficult | 0 | 0 | 0 | 0 | 1 (0.8) |
| *Total* | 172 | 134 | 143 | 154 | 131 |
| **P-value** | p=0.772[2] | | | | |
| **Seeking further help** | | | | | |
| Very easy | 89 (61.8) | 63 (55.8) | 73 (60.8) | 82 (63.1) | 72 (68.6) |
| Easy | 37 (25.7) | 33 (29.2) | 32 (26.7) | 32 (24.6) | 22 (21.0) |
| Neutral | 12 (8.3) | 10 (8.8) | 9 (7.5) | 14 (10.8) | 6 (5.7) |
| Difficult | 6 (4.2) | 5 (4.4) | 5 (4.2) | 2 (1.5) | 5 (4.8) |
| Very difficult | 0 | 2 (1.8) | 1 (0.8) | 0 | 0 |
| *Total* | 144 | 113 | 120 | 130 | 105 |
| **P-value** | p=0.425[2] | | | | |
| **Contacting HCP** | | | | | |
| Very easy | 78 (53.8) | 59 (51.8) | 66 (54.5) | 69 (54.8) | 64 (57.7) |
| Easy | 44 (30.3) | 38 (33.3) | 32 (26.4) | 38 (30.2) | 33 (29.7) |
| Neutral | 14 (9.7) | 11 (9.6) | 11 (9.1) | 16 (12.7) | 6 (5.4) |
| Difficult | 8 (5.5) | 6 (5.3) | 10 (8.3) | 3 (2.4) | 5 (4.5) |
| Very difficult | 1 (0.7) | 0 | 2 (1.7) | 0 | 3 (2.7) |
| *Total* | 136 | 114 | 121 | 126 | 111 |
| **P-value** | p=0.923[2] | | | | |
| **Rating of the service** | | | | | |
| Excellent | 116 (65.2) | 87 (65.9) | 105 (71.4) | 108 (67.5) | 97 (71.9) |
| Good | 53 (29.8) | 40 (30.3) | 36 (24.5) | 40 (25.0) | 29 (21.5) |
| Neutral | 9 (5.1) | 4 (3.0) | 5 (3.4) | 11 (6.9) | 5 (3.7) |
| Poor | 0 | 1 (0.8) | 0 | 1 (0.6) | 3 (2.2) |
| Very poor | 0 | 0 | 1 (0.7) | 0 | 1 (0.7) |
| *Total* | 178 | 132 | 147 | 160 | 135 |
| **P-value** | p=0.699[2] | | | | |

| | | | | | |
|---|---|---|---|---|---|
| **How helpful was the service** | | | | | |
| Very helpful | 113 (64.2) | 83 (64.3) | 98 (67.1) | 97 (61.8) | 91 (68.4) |
| Helpful | 48 (27.3) | 34 (26.4) | 34 (23.3) | 47 (29.9) | 30 (22.6) |
| Neutral | 15 (8.5) | 9 (7.0) | 8 (5.5) | 7 (4.5) | 7 (5.3) |
| Not very helpful | 0 | 2 (1.6) | 4 (2.7) | 4 (2.5) | 3 (2.3) |
| Not at all helpful | 0 | 1 (0.8) | 2 (1.4) | 2 (1.3) | 2 (1.5) |
| *Total* | 176 | 129 | 146 | 157 | 133 |
| **P-value** | p=0.874[2] | | | | |
| **How reassuring was the service** | | | | | |
| Very reassured | 117 (66.1) | 84 (63.2) | 103 (70.1) | 103 (64.8) | 94 (70.1) |
| Reassured | 46 (26.0) | 36 (27.1) | 34 (23.1) | 43 (27.0) | 32 (23.9) |
| Neutral | 14 (7.9) | 10 (7.5) | 6 (4.1) | 8 (5.0) | 4 (3.0) |
| Not very reassured | 0 | 1 (0.8) | 2 (1.4) | 5 (3.1) | 3 (2.2) |
| Not at all reassured | 0 | 2 (1.5) | 2 (1.4) | 0 | 1 (0.7) |
| *Total* | 177 | 133 | 147 | 159 | 134 |
| **P-value** | p=0.612[2] | | | | |

Note. Bold text indicates significance at p< .05 level.
p values are derived from Mann-Whitney U test[1] or Kruskal-Wallis H test[2]
* Deprivation by LSOA (Index of Multiple Deprivation decile)
